# Integrative Analyses Reveal Novel Disease-associated Loci and Genes for Idiopathic Pulmonary Fibrosis

**DOI:** 10.1101/2021.05.11.21257064

**Authors:** Ming Chen, Yiliang Zhang, Taylor S. Adams, Dingjue Ji, Wei Jiang, Louise V. Wain, Michael H. Cho, Naftali Kaminski, Hongyu Zhao

## Abstract

**Background:** Although genome-wide association studies have identified many genomic regions associated with idiopathic pulmonary fibrosis (IPF), the causal genes and functions remain largely unknown. Many bulk and single-cell expression data have become available for IPF, and there is increasing evidence suggesting a shared genetic basis between IPF and other diseases.

**Methods:** By leveraging shared genetic information and transcriptome data, we conducted an integrative analysis to identify novel genes for IPF. We first considered observed phenotypes, polygenic risk scores, and genetic correlations to investigate associations between IPF and other traits in the UK Biobank. We then performed local genetic correlation analysis and cross-tissue transcriptome-wide association analysis (TWAS) to identify IPF genes. We further prioritized genes using bulk and single-cell gene expression data.

**Findings:** We identified 25 traits correlated with IPF on the phenotype level and seven traits genetically correlated with IPF. Using local genetic correlation, we identified 12 candidate genes across 14 genomic regions, including the *POT1* locus (p-value = 4·1E-4), which contained variants with protective effects on lung cancer but increasing IPF risk. Using TWAS, we identified 36 genes, including 12 novel genes for IPF. Annotation-stratified heritability estimation and differential expression analysis of downstream-regulated genes suggested regulatory roles of two candidate genes, *MAFK* and *SMAD2*, on IPF.

**Interpretation:** Our integrative analysis identified new genes for IPF susceptibility and expanded the understanding of the complex genetic architecture of IPF.

**Funding:** NIHR Leicester Biomedical Research Centre, Three Lakes Partners, the National Institutes of Health, the National Science Foundation, U01HL145567, and UH2HL123886.

## Introduction

Idiopathic pulmonary fibrosis (IPF) is a rare and fatal disease[1]. While recently developed therapies mitigate symptoms and slow disease progression, IPF has no cure[2]. In recent years, several common and rare genetic variants, implicating genes involved in alveolar stability, telomere biology, host defense, and cellular barrier function[3, 4], have been associated with IPF. Despite these findings, our understanding of IPF pathogenesis remains elusive. The identification and interpretation of genetic risk factors will facilitate the understanding of molecular mechanisms involved in the pathogenesis of IPF, which could potentially lead to new treatments. However, due to limited sample size, genome-wide association studies (GWAS) have only identified tens of risk loci for IPF[4-6], and the biological interpretations behind GWAS signals remain largely unknown.

Increasing evidence from large GWAS suggests that most trait-associated loci can influence multiple traits. Risk genes such as *TERT, DSP*, and *FAM13A* have been consistently identified for IPF, chronic obstructive pulmonary disease (COPD), and / or lung cancer[7]. Some transcriptomic pathways and metabolite regulations are also shared between COPD and IPF[8, 9]. These findings suggest that novel IPF genetic risk factors could be identified by leveraging shared genetics between traits. Recent developments in multi-trait analysis have led to the emergence of new methods that study the shared genetic basis across multiple phenotypes[10-15]. In particular, genetic correlation[11] and local genetic correlation[14, 15] contribute to a better understanding of shared genetic architecture and pathways between traits. Multi-trait association analysis[12, 16-18], such as multi-trait association mapping (MTAG)[12], can substantially improve GWAS power and facilitate post-GWAS analysis.

Transcriptomic studies provide another resource to identify novel biomarkers and biological interpretation for IPF risk loci. Data generated from large consortium efforts such as the Genotype-Tissue Expression (GTEx) project, ENCODE[19], and IMPACT[20] have provided comprehensive functional annotations for SNPs. Integrating these data can increase power and help biological interpretation of IPF GWAS results and prioritize potential effector genes. For example, transcriptome-wide association studies (TWAS)[21] integrating GWAS results and expression quantitative trait loci (eQTL) information is a powerful tool to bridge SNPs and complex traits through gene expression.

This manuscript aims to identify novel genetic risk factors for IPF through multi-trait modeling and TWAS to overcome the limitations in sample size and statistical power (**Figure 1**). We first investigated the relationship between IPF and other traits through phenotype-level correlation, polygenic risk score (PRS) correlation, and genetic correlation[22]. We then estimated local genetic correlation[15] between IPF and other traits to further identify local regions with shared genetic effects and prioritized 12 candidate genes from the identified regions. In addition, we performed cross-tissue expression imputation and gene-level association analysis to identify 12 additional candidate risk genes[23]. To validate our findings, we examined the expression patterns of candidate genes in bulk and single-cell expression data[24, 25]. In particular, we demonstrated the regulatory role of two transcription factors (TFs), *MAFK* and *SMAD2*, through heritability enrichment analysis and cell-type-specific differently expressed gene enrichment analysis of their target genes.

**Figure 1.**
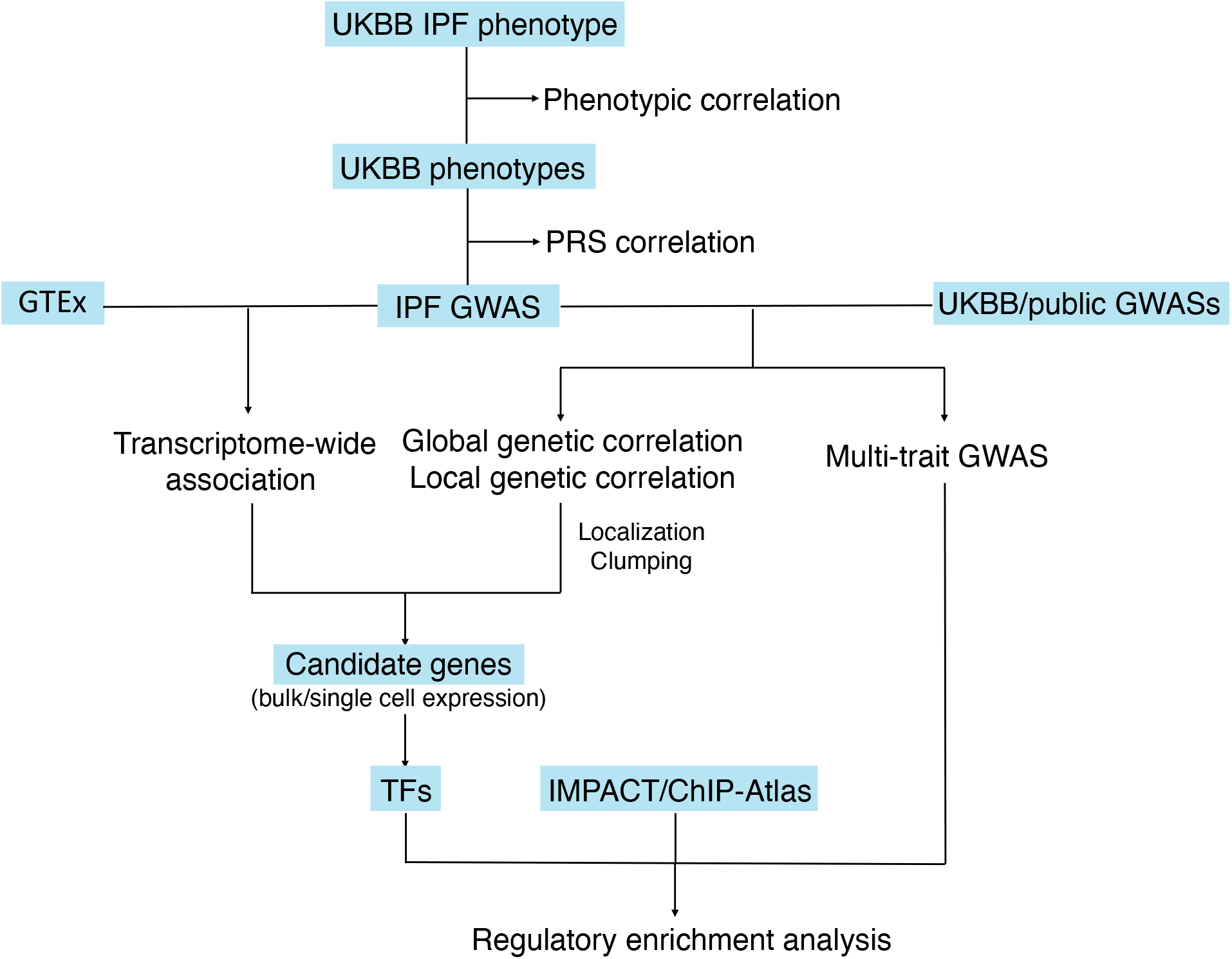
The integrative analysis workflow for candidate IPF genes identification. Abbreviations used are: UKBB (UK Biobank) [26], GTEx (The Genotype-Tissue Expression project) [30], GWAS (genome-wide association study), TF (transcription factor). IMPACT is a genomic annotation tool of cell-state-specific regulatory elements inferred from the epigenome of bound transcription factors [32]. ChIP-Atlas is a public database for ChIP-seq data [35].

## Methods

### Phenotypic correlation

We investigated the phenotype-level correlation between IPF and phenotypes in the UK Biobank (UKBB)[26]. The definition of IPF cases was a combination of data field 22135 (Doctor diagnosed idiopathic pulmonary fibrosis) and data field 41202 (Diagnoses - main ICD10) with ICD10 code J84 (Pulmonary fibrosis, unspecified) in UKBB. For the other traits in UKBB, each trait corresponds to one data field in UKBB. We used only baseline measurements (UKBB UDI codes end with 0.0) and filtered out a) traits with <1000 observations, b) repetitive or highly similar traits, c) traits reflected transient features (e.g., recent diet). We applied logistic regression and McFadden pseudo-R-square coefficients[27], a measure to evaluate the goodness-of-fit of logistic models, to obtain phenotype-level correlations (FDR < 0·05). We ultimately arrived at a total of 670 phenotypes with estimated phenotype-level correlations.

### Polygenic risk score correlation

Besides phenotype-level correlation, we also investigated the relationship between a polygenic risk score (PRS) of IPF and the 670 phenotypes in UKBB. We used the R package EB-PRS[28] to obtain PRS for individuals from the UKBB based on IPF summary statistics[4]. Following standard quality control criteria, we restricted the analysis to autosomal variants with genotype missing rate per marker < 0·05, imputation information score above 0·3, Hardy-Weinberg Equilibrium *p*-value > 1e-4, and minor allele frequency (MAF) < 0·01. We further removed individuals by considering the following criteria: non-British, relatedness, sex aneuploidy, discordant sex, heterozygosity, missingness, and kinship. As a result, 226,736 individuals were removed. The 275,896 remaining individuals were used for PRS calculation. Then we calculated the correlation between IPF PRS and the 670 UKBB traits. Linear regression, logistic regression, and ordinal/multinomial regression were used to calculate the correlation between PRS of IPF for continuous traits, binary traits, and categorical traits, respectively. We calculated R-square coefficients for continuous traits and McFadden pseudo-R-square[27] for the other traits (FDR < 0·05).

### Genetic correlation

We examined genetic correlation using the IPF GWAS and other phenotypes. GWAS summary statistics were from 1) UKBB and 2) non-UKBB. For the UKBB, we obtained GWAS summary statistics from the 2nd round results (**URLs**). We manually removed repeated and similarly defined traits, and 350 traits remained for analysis. To better match the phenotypes used in previous analyses but not available from UKBB GWAS results, we downloaded summary statistics for 31 traits from publicly available GWAS results. All the GWASs were performed on samples majorly from European ancestry. Details, including the sample sizes and the resources of the GWASs, are summarized in the **Supplementary file**. To investigate the genetic similarity between IPF and other traits, we estimated the genetic correlations based on GWAS summary data using the software GNOVA[22].

### Local genetic correlation and prioritization of candidate genes

To improve the stability of local genetic correlation estimation, we selected 17 traits that had absolute values of genetic covariance with IPF greater than 0·02 and FDR < 0·05 from our genetic correlation results. Local genetic correlations were calculated using the gene covariance analysis software SUPERGNOVA (**URLs**)[15]. To quantify the degree of local correlation, we estimated the proportion of correlated regions with the R package *ashr*[29]. The inputs were estimates of local genetic covariance and their standard errors. The unimodal prior distribution was set to be “halfnormal” for all the results of pairs of traits. The method applied a Bayesian framework to compute FDR for each genomic region. To estimate the number of correlated regions for each pair of traits, we calculated the sum of (1 – FDR) given by *ashr* for each region. To prioritize genes with potential pleiotropic effects on IPF and the other trait on local regions, we first clumped the SNPs in the significantly correlated regions using the genome analysis software PLINK (**URLs**). We set the significance threshold for index SNPs as 0·001 and 0·01, linkage disequilibrium (LD) threshold for clumping as 0·2, and the physical distance threshold for clumping as 250 kb for all regions. After clumping, the variant with the lowest *p*-value in each region was defined as the sentinel variant. Next, the sentinel variants were mapped to the gene or the nearest gene.

### Transcriptome-wide association study (TWAS)

We used a joint-tissue TWAS method called UTMOST (**URLs**)[23] to identify IPF-associated genes with its pre-trained built-in gene expression imputation model using 44 tissues from GTEx[30]. The associations between phenotype and imputed gene expression level across tissues were jointly tested to improve the power. Bonferroni correction was used to adjust for multiple hypothesis testing. We further applied conditional analysis to prioritize candidate genes located within 1 million base pairs.

### Multi-trait analysis and partitioned heritability

To further explore TFs from our candidate genes, we applied LDSC (**URLs**)[31] to investigate the annotation-stratified heritability enrichment of each TF on IPF GWAS summary statistics[4]. The input SNP annotations were defined by the TF binding sites in various cell types from IMPACT [32]. We conditioned the analysis on the 52 baseline annotations in LDSC. The same procedure mentioned above was also performed on the IPF GWAS summary statistics adjusted by MTAG to boost the power of the original IPF GWAS summary statistics. For MTAG, we selected traits that had estimated heritability > 0·2, the absolute value of genetic covariance with IPF > 0·02, and FDR < 0·05 from our genetic correlation results. In the end, four traits, including whole-body fat mass, body fat percentage, arm fat percentage, and hip circumference, were used for pair-wise MTAG with IPF.

### Bulk and single-cell expression analysis

For bulk expression data, we obtained curated expression data of IPF and controls from the PulmonDB[25] web-based database (**URLs**). The studies used were GSE21369, GSE26594, GSE31934, GSE32537, GSE35145, GSE38958, GSE44723, GSE45686, GSE48149, GSE52463, GSE53845, GSE6804, GSE71351, GSE72073. PulmonDB provides the log-ratio between the expression value in test and reference conditions. We extracted the log-ratio for genes annotated as “IPF” as the test and “HEALTHY_CONTROL” or “MATCH_TISSUE_CONTROL” as reference. To determine DEGs, we conducted a t-test for each candidate gene in each study with a significance level of 0·05 after Bonferroni correction. For single-cell expression analysis, single-cell expression data for 32 IPF cases and 28 healthy controls were obtained from the IPF Cell Atlas[24]. Cell-type-specific DEGs were obtained using the MAST (Model-based Analysis of Single-cell Transcriptomics)[33] test implemented in the R package of Seurat (**URL**)[34]. To test the enrichment of TF targets in DEGs for each cell type, we applied the hypergeometric test using cell-type-specific DEGs, and TF target gene sets (**Supplementary file**) predicted from the ChIP-Atlas(**URLs**)[35, 36]. We investigated the proportions of cells expressing candidate genes using two-proportions z-test and adjusted p-value with Bonferroni correction.

## Results

### Analytic strategies

To increase the statistical power of current IPF GWAS and facilitate fine-mapping of identified GWAS signals, we first scanned for IPF-related traits through phenotype-level correlations, polygenic risk score correlations, and genetic correlations. For genetic correlation, we examined global genetic correlation and local genetic correlation to identify genetically correlated traits and understand regional pleiotropic effects. For local correlated regions, we localized the signals to prioritize candidate genes. Second, we used TWAS to identify genes whose expression is genetically correlated with IPF. Furthermore, to validate TWAS and multi-trait analysis results, we used the multi-trait GWAS method to increase IPF GWAS power by leveraging the pleiotropic effect. We examined the partitioned heritability enrichment of identified transcription factors. We also examined the downstream gene expression with IPF single-cell sequencing data. **Figure 1** shows the overall workflow of this study.

### Phenotype-level correlation, PRS correlation, and genetic correlation between IPF and UK Biobank traits

While the correlation strength between IPF and other traits was weak in general, 25 traits had significant phenotype-level correlations with IPF after Bonferroni correction (**Figure 2A; Supplementary file**). Top correlated phenotypes are related to complex diseases, e.g., Systemic Lupus Erythematosus (coef = 4·46, adjusted *p*-value = 2·27E-15) and Doctor diagnosed emphysema (coef = 3·44, adjusted *p*-value = 5·51E-15), hematological traits, e.g., lymphocyte percentage (coef = -8·7E-04, adjusted *p*-value = 4·39E-05), age and overall health, e.g., number of treatments/medications taken (coef = 0·18, adjusted *p*-value = 3·16E-11) and age at recruitment (coef = 0·12, adjusted *p*-value = 2·20E-07).

**Figure 2.**
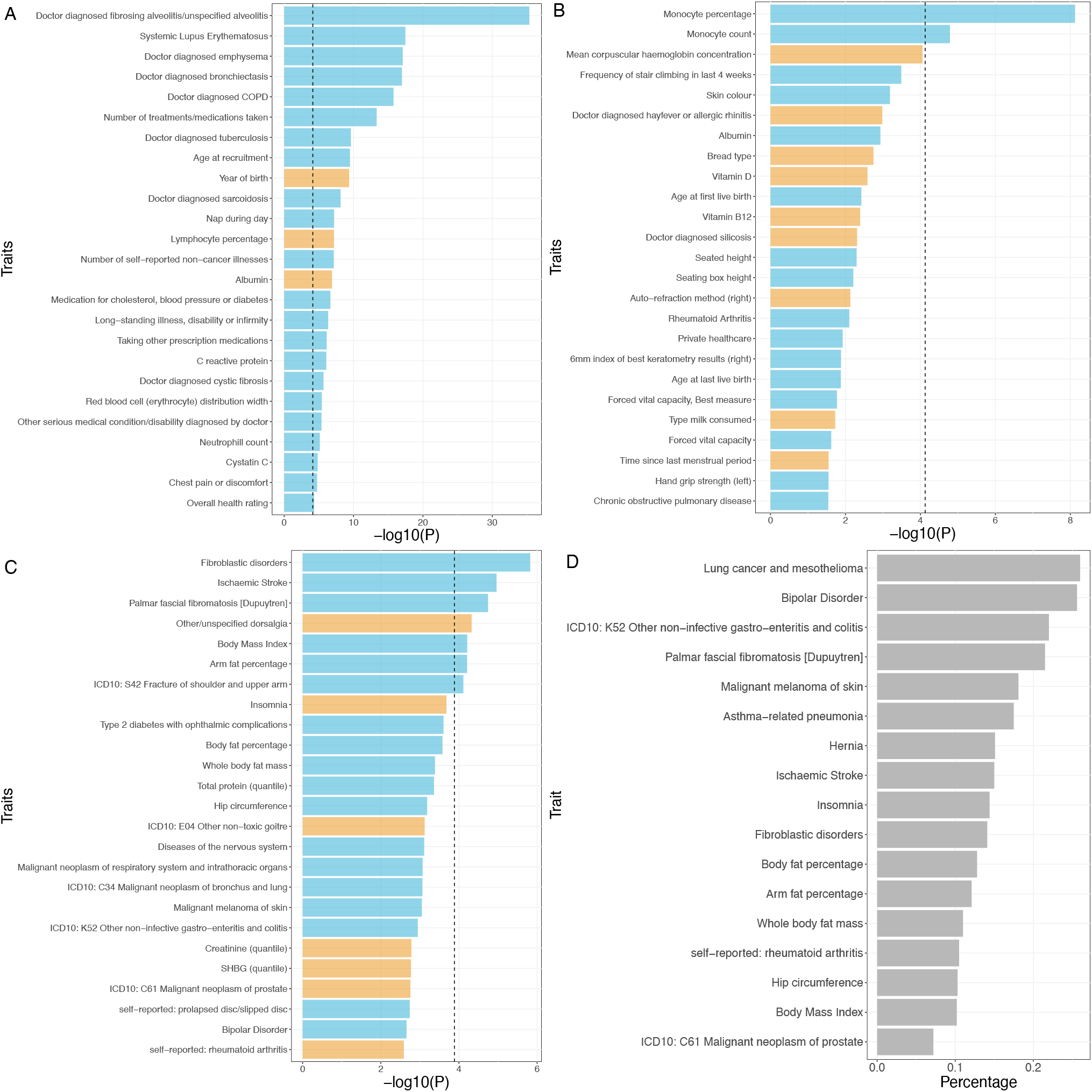
Bar plots of correlations between IPF and UK Biobank phenotypes. The black dashed line corresponds to the Bonferroni adjusted p-value = 0·05. Positive and negative correlations are highlighted in blue and yellow, respectively. Top 25 UK Biobank traits ranked by the adjusted p-value of the (A) phenotype-level correlations with IPF. (B) polygenic risk score correlations with IPF. (C) global genetic correlation with IPF. (D) Bar plot of the proportions of correlated local regions between IPF and 17 complex traits with significant global correlations with IPF and the absolute value of genetic covariance > 0·2.

We use PRS correlation to understand how the genetic risk of IPF is related to other phenotypes (**Figure 2B; Supplementary file**). We identified two traits with significant correlations. They are monocyte percentage (coef = 1·69E-10, adjusted *p*-value = 4·99E-06) and monocyte count (coef = 1·51E-09, adjusted *p*-value = 0·011). Notably, a significant positive correlation between IPF mortality and monocyte percentage was observed in clinical studies, suggesting monocyte count/percentage as a potential prognostic marker for IPF [26, 27].

To understand the shared genetic etiology, we identified seven traits having significant genetic correlations with IPF after Bonferroni correction (**Figure 2C; Supplementary file**). Similar to phenotype-level correlation, the genetic correlation between IPF and other traits was weak in general. Top correlated traits are related to connective tissue diseases, e.g., fibroblastic disorders (rho = 0·027, adjusted *p*-value = 0·00057) and other/unspecified dorsalgia (rho = -0·016, adjusted *p*-value = 0·018), body fat, e.g., Body Mass Index (rho = 0·024, adjusted *p*-value = 0·023).

To improve the resolution of genetic correlation, we calculated the local genetic correlation between IPF with 17 top genetically correlated traits (**Methods; Figure 2C**). Among these traits, 14 showed significant local region correlations across 14 local regions (FDR < 0·05; **Supplementary file**). Similar patterns and signals of GWAS marginal statistics are shown in local regions with significant local genetic correlations (**Supplementary file**). We estimated the proportion of correlated regions among whole-genome regions for each trait based on local genetic correlation analysis (**Figure 2D; Methods**). Although the global genetic correlation between IPF and lung cancer only ranks middle among the 17 traits in terms of both the correlation and significance level (cov = 0·014, adjusted *p*-value = 0·019), it has the largest proportion of correlated regions (26% for squamous cell lung cancer). In contrast, the traits showing strong global correlations with IPF, such as fibroblastic disorders and ischemic stroke (adjusted *p*-value= 0·00057 and 0·0042, respectively), only have a moderate proportion of correlated regions (14. 1% and 15%, respectively). In addition, we found that similar phenotypes were more likely to be correlated with IPF at the same genomic regions (**Supplementary file**). For example, there was significant local genetic correlation at a region (chr8:108,646,968-110,761,074) between IPF and fibroblastic disorders (cov = 0·0015, adjusted *p*-value = 2·1E-04) and palmar fascial fibromatosis (cov = 0·0014, adjusted *p*-value = 2·1E-04).

### Local genetic correlation and TWAS identified candidate IPF risk factors

Local genetic correlation analysis helped to identify local genomic regions/genes with pleiotropic effects. Notably, all of the significantly correlated regions did not harbor any SNPs reaching genome-wide significance for IPF. Therefore, genes in these regions are less likely to be captured in the post-GWAS analysis. For example, we identified significant local genetic correlation in a region (chr4:145,024,452-148,047,972; **Supplementary file**) for hip circumference [28, 29]. However, the most significant *p*-value for IPF of the SNPs in that region is 7·8E-4 (rs2055059). These findings motivated us to further fine-map local pleiotropic effects and find candidate pleiotropic genes. Thus, we refined association signals on the local regions and identified 12 candidate genes (**Table 1; Methods)**. Many of these genes have been reported to be either directly related to IPF or IPF related pathogenic functions like telomere maintenance and TGF-β signaling.

**Table 1.**
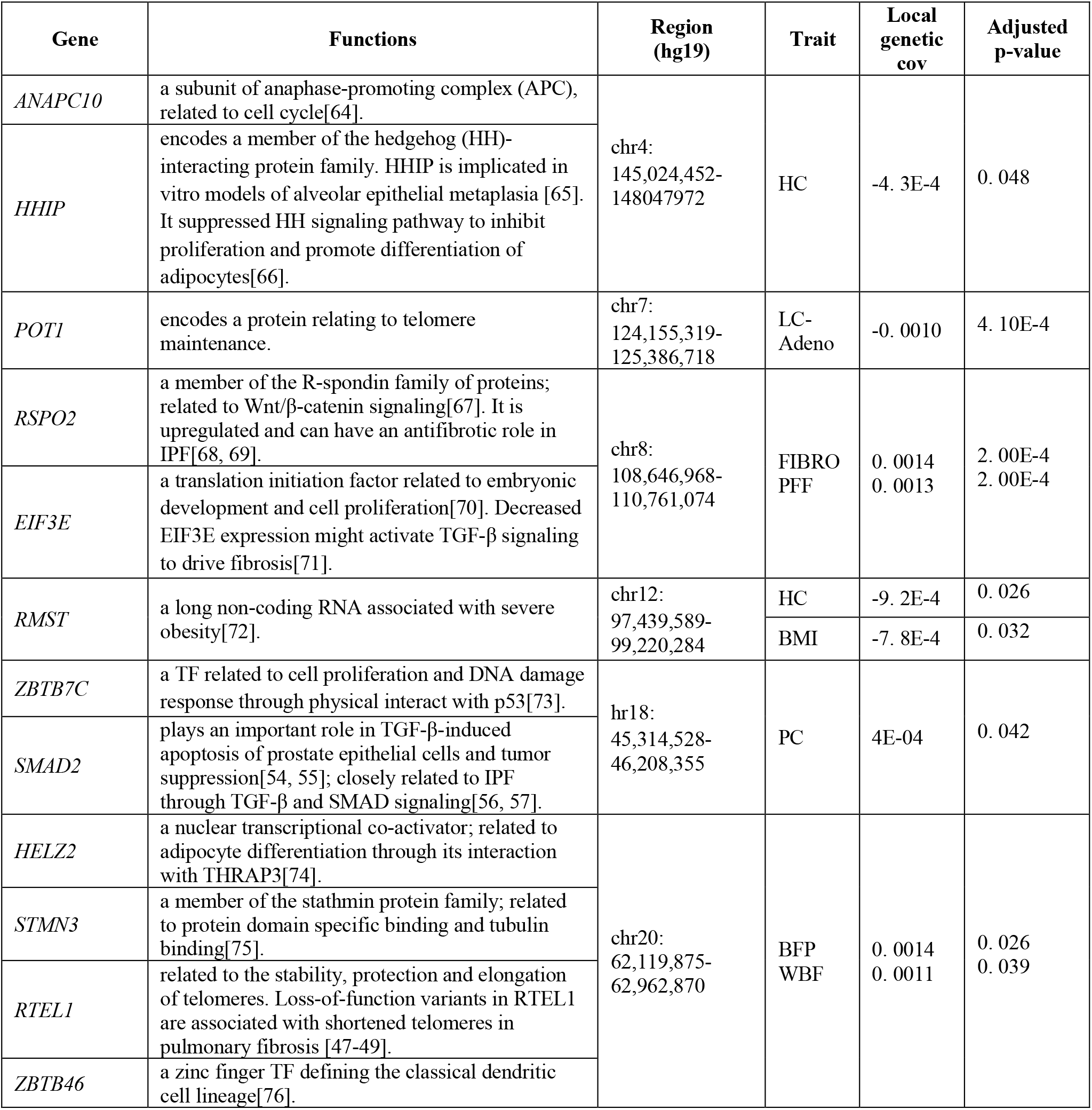
Novel IPF candidate genes identified in correlated local regions. (LC-ever: lung cancer (ever smoking); LC-Adeno: lung cancer (adenocarcinoma); HC: hip circumference; FIBRO: fibroblastic disorders; PFF: palmar fascial fibromatosis; PC: malignant neoplasm of prostate; BFP: body fat percentage; WBF: whole-body fat mass; BMI: body mass index)

| Gene | Functions | Region (hg19) | Trait | Local genetic cov | Adjusted p-value |
| --- | --- | --- | --- | --- | --- |
| *ANAPC10* | a subunit of anaphase-promoting complex (APC), related to cell cycle[64]. | chr4:<br>145,024,452-148047972 | HC | -4. 3E-4 | 0. 048 |
| *HHIP* | encodes a member of the hedgehog (HH)-interacting protein family. HHIP is implicated in vitro models of alveolar epithelial metaplasia [65]. It suppressed HH signaling pathway to inhibit proliferation and promote differentiation of adipocytes[66]. |  |  |  |  |
| *POT1* | encodes a protein relating to telomere maintenance. | chr7:<br>124,155,319-125,386,718 | LC-Adeno | -0. 0010 | 4. 10E-4 |
| *RSPO2* | a member of the R-spondin family of proteins; related to Wnt/β-catenin signaling[67]. It is upregulated and can have an antifibrotic role in IPF[68, 69]. | chr8:<br>108,646,968-110,761,074 | FIBRO PFF | 0. 0014<br>0. 0013 | 2. 00E-4<br>2. 00E-4 |
| *EIF3E* | a translation initiation factor related to embryonic development and cell proliferation[70]. Decreased EIF3E expression might activate TGF-β signaling to drive fibrosis[71]. | | | | |
| *RMST* | a long non-coding RNA associated with severe obesity[72]. | chr12:<br>97,439,589-99,220,284 | HC | -9. 2E-4 | 0. 026 |
|  |  |  | BMI | -7. 8E-4 | 0. 032 |
| *ZBTB7C* | a TF related to cell proliferation and DNA damage response through physical interact with p53[73]. | chr18:<br>45,314,528-46,208,355 | PC | 4E-04 | 0. 042 |
| *SMAD2* | plays an important role in TGF-β-induced apoptosis of prostate epithelial cells and tumor suppression[54, 55]; closely related to IPF through TGF-β and SMAD signaling[56, 57]. | | | | |
| *HELZ2* | a nuclear transcriptional co-activator; related to adipocyte differentiation through its interaction with THRAP3[74]. | chr20:<br>62,119,875-62,962,870 | BFP WBF | 0. 0014<br>0. 0011 | 0. 026<br>0. 039 |
| *STMN3* | a member of the stathmin protein family; related to protein domain specific binding and tubulin binding[75]. |  |  |  |  |
| *RTEL1* | related to the stability, protection and elongation of telomeres. Loss-of-function variants in RTEL1 are associated with shortened telomeres in pulmonary fibrosis [47-49]. |  |  |  |  |
| *ZBTB46* | a zinc finger TF defining the classical dendritic cell lineage[76]. |  |  |  |  |

TWAS incorporate eQTL information of genes to improve the statistical power and biological interpretability of GWAS results. Thirty-seven genes were identified as significant for IPF through the UTMOST TWAS test using 44 GTEx tissues after Bonferroni correction, and 36 of these genes remained significant after conditional analysis (**Supplementary file**). Of these genes, 24 of them have been reported in IPF GWAS, TWAS, or found to be differentially expressed in IPF patients versus healthy individuals [4, 8, 30-35]. Among the 12 newly identified genes for IPF, four are in different risk loci from the other 24 genes (**Supplementary file**). The detailed information for these 12 genes can be found in **Table 2**.

**Table 2.** Novel IPF candidate genes that were identified in the transcriptome-wide association study.

| Gene | Functions | Position(hg19) | Adjusted p-value |
| --- | --- | --- | --- |
| *PAK6* | Inhibition of *PAK6* led to reduction in cell proliferation, migration and invasion of the cigarette smoke treated cells [77]. | chr4:40,509,629-40,569,688 | 1. 35E-2 |
| *MAFK* | a TF belonging to the small Maf proteins (sMaf). The Bach1-sMaf heterodimer regulates the heme oxygenase-1 (*HMOX1*) gene [73], which was found to be a down-regulated DEG of IPF [74]. | chr7:1,570,350-1,582,679 | 2. 00E-2 |
| *HRAS* | belongs to the Ras oncogene family; *HRAS* mutations were detected in patients having lung cancer with IPF[78]. | chr11:532,242-537,287 | 2. 62E-02 |
| *LMNTD2* | a protein coding gene. | chr11:554,850-560,779 | 2. 75E-02 |
| *CDHR5* | a novel mucin-like gene that is a member of the cadherin superfamily. Associated with many metabolic and some tumor growth-associated processes and pathways[79]. | chr11:616,565-626,078 | 1. 70E-07 |
| *SCT* | encodes a member of the glucagon family of peptides. | chr11:626,313-627,173 | 4. 26E-06 |
| *SLC25A22* | encodes a mitochondrial glutamate carrier. | chr11:790,475-798,316 | 9. 21E-06 |
| *PRR33* | a protein coding gene. | chr11:1,910,375-1,912,084 | 7. 72E-6 |
| *TNNT3* | related to skeletal and muscular system development and function [80]. | chr11:1,940,792-1,959,936 | 2. 56E-6 |
| *BMF* | belongs to the BCL2 protein family; act as an important mediator of cell death signaling pathways [81]. | chr15:40,380,091-40,401,093 | 7. 14E-6 |
| *GJC1* | a protein component of gap junction; GJC1 hypermethylation was seen among a small subset of adenomas [82]. | chr17:42,875,816-42,908,184 | 4. 24E-6 |
| *ARL17A* | encodes a protein of the ADP-ribosylation factor family; involves in multiple regulatory pathways relevant to human carcinogenesis [83]. | chr17:43,697,710-43,913,194 | 4. 62E-3 |

Altogether we have 24 new candidate genes, 12 of them were obtained from local genetic correlation, and 12 were obtained using TWAS. Next, we investigated their expression using both bulk and single-cell expression data. From bulk expression, six genes *CDHR5, LMNTD2, SLC25A22, HRAS, SCT, POT1*(**Figure 3A; Supplementary file**) in total were found to be differentially expressed in IPF patients verse healthy controls (*p*-value < 0·05/24). In single-cell data, we found *HHIP* (logFC = -0·41, adjusted *p*-value = 2·67E-28) and *RMST* (logFC = -0·41, adjusted *p*-value = 2·67E-28) differentially expressed between IPF and healthy individuals in ATII cell type. 17 out of 24 genes have a higher proportion of cells expressing them in IPF patients than control individuals (**Figure 3B**).

**Figure 3.**
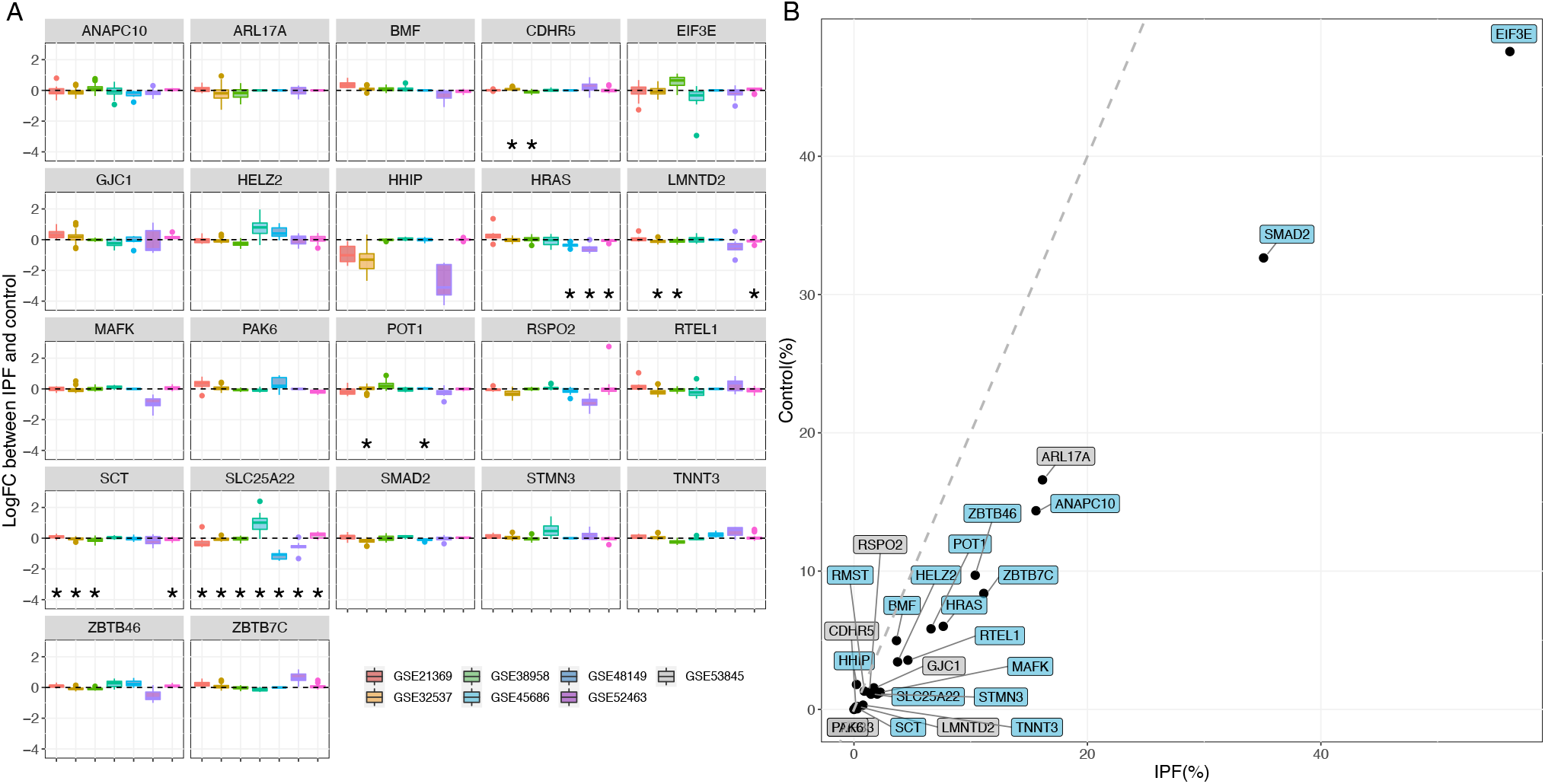
Expression patterns of 24 IPF candidate genes. (A) Boxplot of bulk expression of candidate genes across seven studies. Differentially expressed genes labeled with black asterisk sign were obtained using T-test after Bonferroni correction with adjusted p-value < 0·05. *PAK6* and *PRR33* are not shown here because they were not present in the bulk expression data. (B) Cell proportions of expressing candidate genes in IPF patients compared to healthy individuals in single-cell data. Genes with significantly different proportions are highlighted in blue (two-proportions z-test with adjusted p-value < 0·05 after Bonferroni correction). Thegrey dashed line represents y = x.

### The regulatory role of candidate TFs in IPF

There are seven TFs among the 24 identified candidate genes, including *BAHD1, EIF3E, HELZ2, MAFK, SMAD2, ZBTB7C*, and *ZBTB46. MAFK* was identified using TWAS, and the rest TFs were identified by local genetic correlation. Their regulatory roles in IPF are of particular interest. For these seven TFs, we screened their target genes in the genomic annotation data we use. For IMPACT, only *MAFK* has predicted TFBSs in six cell lines. For ChIP-Atlas, *MAFK* has 13 datasets across eight cell lines, and *SMAD2* has 39 datasets across 12 cell lines. The binding site information of the other TFs is unavailable. Therefore, we conducted partitioned heritability analysis for *MAFK* and enrichment analysis for *MAFK* and *SMAD2* target genes.

The partitioned IPF heritability showed moderate enrichment for the TFBSs of *MAFK* but failed to reach a significant level after adjusting multiple tests **(Supplementary file**). To boost the statistical power of partitioned heritability analysis through joint analysis, we applied MTAG [1] on GWAS summary statistics of IPF with 17 top genetically correlated traits (**Methods, Figure 2C**). The partitioned heritability of *MAFK* TFBSs after joint analysis had significant enrichment on all eight cell types, including the cervix cell, stem cell, liver cell, lung cell from fibroblasts, myeloid and B cell from the blood (**Figure 4A; Supplementary file**). Partitioned heritability of *MAFK* TFBS in lung cells from fibroblasts consistently has the most significant enrichment results across the four MTAG-ed GWAS summary data (Supplementary file), indicating that the regulatory effect of *MAFK* is most significant in IPF disease-relevant tissue. Furthermore, to ensure the change of the enrichment analysis results were not artifacts due to the effect of the auxiliary traits used in MTAG, we conducted the same partitioned heritability analysis for the traits used in MTAG alone, i.e., hip circumference and whole-body fat mass, and none showed significant enrichment for the TFBSs of *MAFK* in any cell type (**Supplementary file**).

**Figure 4.**
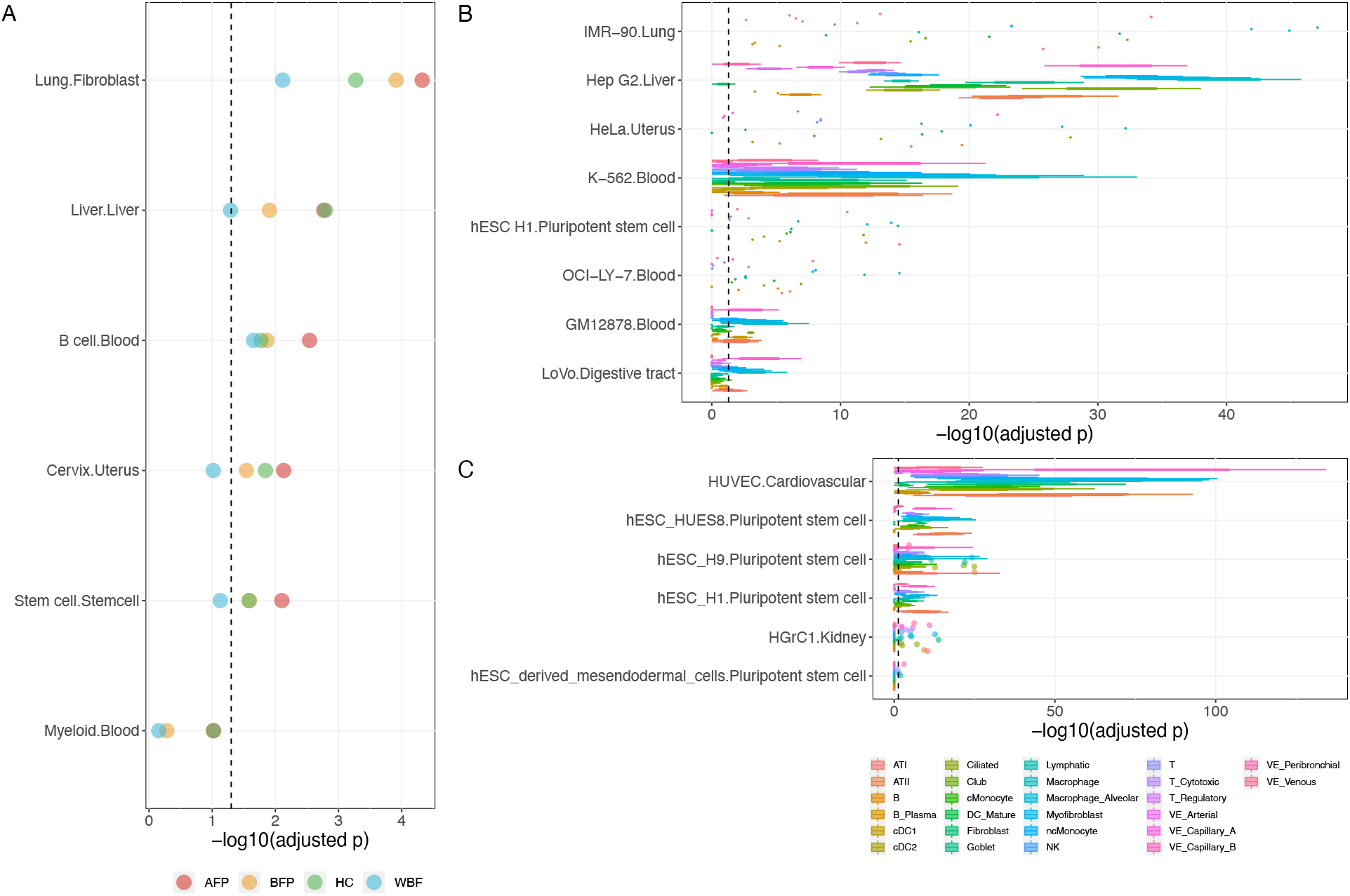
Regulatory enrichment analysis of *MAFK* and *SMAD2*. The black dashed line corresponds to the Bonferroni adjusted p-value = 0·05. (A) Partitioned heritability enrichment analysis using the IPF GWAS summary statistics data after MTAG multi-trait analysis. Y-axis represents different annotations of predicted TF binding sites of *MAFK* in six cell types from the IMPACT study. For each annotation, dots represent p-values of the hypergeometric test result and are colored by traits used for MTAG. (B) Boxplot for enrichment analysis of *MAFK* regulated genes among cell-type-specific DEGs between IPF patients and healthy individuals. Y-axis is cell-state-specific annotations of *MAFK* target genes from ChIP-Atlas. One cell-state-specific annotation can have multiple sample sources. For each annotation, boxplots represent sample hypergeometric test p-values and are colored by cell type. Only annotations and cell-types with at least one significant enrichment result are plotted here. (C) Similar boxplot as (b) for *SMAD2*.

We then examined the enrichment of *MAFK* and *SMAD2* target genes within cell-type-specific differentially expressed genes (DEGs) between IPF patients and healthy individuals using single-cell data. For *MAFK*, all of its targeted gene sets showed significant enrichment in cell-type-specific DEGs after Bonferroni correction across the eight cell lines (**Figure 4B; Supplementary file**). The top enrichments were mostly among myofibroblast, macrophage, and alveolar macrophage for the annotations in the IMR-90 cell line from the lung. For *SMAD2*, 6 out of 12 targeted gene sets had significant enrichment in cell-type-specific DEGs after Bonferroni correction (**Figure 4C; Supplementary file 9**). The top enrichments were mostly among vascular endothelial capillary B cell, myofibroblast, and alveolar macrophage cell for annotations in HUVEC cell line, a cardiovascular cell type obtained from the umbilical cord.

## Discussion

Although previous literature has discussed the relationship between IPF and other lung diseases[7-9, 37], there has not been a comprehensive study of the similarities and differences between IPF and other traits. Using the large-scale biobank data and genomic regulatory information, we have conducted detailed analyses on the between-trait relationship of IPF from different perspectives, including phenotype-level correlation, genetic risk score correlation, and genetic correlation, to understand the degree of pleiotropy. We also employed local genetic correlation and TWAS to identify candidate IPF risk genes to facilitate future research on disease mechanisms and drug development.

First, the between-trait correlation analyses deepened our understanding of the relationship between IPF and other phenotypes. In general, the correlations were weak using different measurements. We found that many top correlated traits are related to fibrosis, malignant neoplasm, and immune diseases, suggesting possible mechanisms of IPF overlapping with those phenotypes. For lung-related phenotypes, although asthma, COPD, and decreased lung functions have significant phenotype-level correlations with IPF, their genetic correlations are not apparent. We did not find any significant correlations with smoking behaviors. In addition, many correlated traits are associated with aging, such as overall health conditions, obesity, and cardiovascular diseases. Especially, we found significant genetic correlations between IPF and many body fat related traits. Different studies have found that obesity is a common comorbidity of IPF[38, 39]. Lower body mass index (BMI) and body weight loss seemed to be related to poor outcomes [40]. Altogether, the convergence of genetic and phenotype-level results suggested that metabolic dysregulation might be a critical contributor to the pathogenesis of IPF[41]. Notably, many top correlated phenotypes such as BMI, emphysema, FVC, monocytes, albumin, and neutrophilia appeared to be key IPF survival predictors or clinical markers[42]. In the future, it will be worthwhile to study other correlated phenotypes identified in this study to understand their clinical applications.

We used local genetic correlation analysis to improve the resolution of the pleiotropy between complex traits. We provided a novel angle to find risk genes by leveraging the local pleiotropic effect. The results may provide insights for studying the pathway and mechanisms shared between diseases and suggest possible treatments under comorbidities. For example, *POT1, ZBTB7C*, and *SMAD2* were found to be shared between IPF and cancers. *POT1* was identified in region chr7:124,155,319-125,386,718 for lung cancer. *ZBTB7C* and *SMAD2* was identified in region chr18:45,314,528-46,208,355 for malignant neoplasm of prostate. IPF and cancer share many common risk factors, and both lung cancer and prostate cancer have higher incidence rates among IPF patients[43]. *POT1* is involved in telomere maintenance[44, 45]. In a recent GWAS in a Japanese population, *POT1* was associated with lung cancer[46]. Pulmonary fibrosis patients carrying *POT1* variants had shorter telomeres, which led to a worse outcome of IPF[47-49]. This suggests that *POT1* variants might be involved in the process of shortened telomeres and lead to worse outcomes in IPF. In a separate study, patients with high *POT1* expression levels in lung cancer tissues showed an overall better survival rate, indicating a protective role of *POT1* in lung cancer prognosis[50]. These findings showed that telomere length regulation involving *POT1* is a shared disease mechanism of IPF and lung cancer.

*ZBTB7C* is a TF that can interact with p53. It is also related to cell proliferation through glutamine metabolism[51]. Glutamine metabolism was showed to be essential for collagen protein synthesis in lung fibroblasts[52]. Glutamine metabolism was also found to differ in prostate cancer[53]. These results suggested glutamine metabolism involving *ZBTB7C* might be a shared mechanism between prostate cancer and IPF. *SMAD2* plays an important role in TGF-β-induced apoptosis of prostate epithelial cells and tumor suppression[54, 55]. It is closely related to IPF through TGF-β and SMAD signaling to promote extracellular matrix gene expression and fibrosis[56, 57]. These findings suggest that *SMAD2* variants confer similar susceptibility to IPF and lung cancer and might be mediated through the TGF-β signaling. In addition, we found that its target genes have significant enrichment in IPF cell-type-specific DEGs. To conclude, further work is needed to elucidate the roles of the above genes and mechanisms play in IPF and cancers.

Through TWAS, we identified 12 novel genes that were not reported in previous studies. Specifically, *HRAS, SCT, and SLC25A22* are expressed in a higher proportion in IPF cells. They also exhibited positive effects in TWAS in lung tissue. Nevertheless, the directions of their expression regulation vary in different studies in bulk expression. Therefore, we did not see any concordance between eQTL effects and expression level since the absolute proportions of cells expressing these genes are small, and their fold change between IPF patients and control individuals was also small. Furthermore, focused studies with a larger sample size could help investigate the effect of these genes on IPF.

With the help of IMPACT[32], we investigated the regulatory role of the identified TF, *MAFK. MAFK* expressed significantly in a higher proportion of cells in IPF patients. We found significant heritability enrichment on *MAFK* TFBSs, and significant enrichment of *MAFK* targeted genes in DEGs among most cell types in IPF single-cell data. *MAFK* can form a heterodimer to regulate antioxidant and xenobiotic-metabolizing enzyme genes[58]. Studies have identified that *MAFK* can modulate NF-kB activity[59] and can be induced by transforming growth factor-b (TGF-b) to regulate downstream genes[60]. *HMOX1*, regulated by *MAFK*, was found to play a central role in the defense against oxidative and inflammatory insults in the lung[61]] and related to many pulmonary diseases, including asthma, COPD, and IPF. Another downstream gene, *GPNMB*[62, 63], is related to fibrosis by inducing epithelial-mesenchymal transition. These findings suggest that *MAFK* may participate in the pathogenesis of IPF through its relationship with both fibrosis and inflammatory-related processes. We also noticed that some well-known IPF genes like *MUC5B* or *TOLLIP* were not identified in TWAS. The reason was that these genes were not well-imputed because of the models and eQTL data used. Thus they did not appear in the results.

Despite the above exciting novel findings, our study has several limitations. First, we manually selected traits for correlation analysis in an attempt to avoid spurious results. The selection criteria, mixed with a certain degree of subjectivity, might influence the results. Large studies that include sufficient samples of other important diseases may not be found in population-based samples like UK Biobank. Second, some of the top-ranked phenotypes could be cases of misdiagnosis or misclassification or be influenced by their prevalence. For example, Systemic Lupus Erythematosus does not usually result in ILD; asbestosis usually results in UIP but is not idiopathic. Third, although our results suggest local regions may contain disease risk genes that failed to be identified in GWAS, these regions might be false positives because local genetic correlation has lower power and less stable estimation than the global genetic correlation.

Although we pre-filtered regions with a small number of SNPs to improve the local genetic correlation results’ credibility, a better way to prove the relevance of these regions is to replicate the results on an independent dataset. However, such a study with little sample overlap and large sample size is not available currently. In the future, additional studies are needed to investigate whether these candidate genes are genuinely related to IPF.

Taken together, through the investigation of the plethora of datasets, we identified 25 traits with significant phenotype-level correlations and seven traits with significant genetic correlation with IPF. By integrating GWAS data with pleiotropy information and transcriptome data, we discovered 12 genes from local multi-trait results and 12 novel genes from TWAS results. Follow-up analyses showed the differential expression and gene expression regulatory function among these candidate genes. These findings and their implications will provide new avenues for investigating the underlying biology and potential therapeutics in this deadly disease.

## Data Availability

We gratefully acknowledge all the studies and databases that made GWAS summary data, expression data, and annotation data available as listed in the supplemental file. This research was conducted by using the UK Biobank resource under application numbers 29900. All data related to results can be found in the result section and the supplementary file.

## URLs

LDSC (https://github.com/bulik/ldsc)

UK Biobank GWAS data (http://www.nealelab.is/uk-biobank)

SUPERGNOVA (https://github.com/qlu-lab/SUPERGNOVA)

UTMOST (https://github.com/Joker-Jerome/UTMOST/)

Seurat (https://satijalab.org/seurat/)

LocusZoom (http://locuszoom.org/)

PLINK (http://pngu.mgh.harvard.edu/purcell/plink/)

PulmonDB(http://pulmondb.liigh.unam.mx/)

## Contributors

M.C. and Y.Z. contributed to study concept and design, data analysis, and manuscript writing. T.S.A. contributed to IPF single cell data processing, and manuscript writing. D.J. and W.J. contributed to data analysis. L.V.W. and M.H.C provided critical interpretation of the findings. N.K. and H.Z. contributed to study concept, design, and statistical support. All authors contributed to review and editing of the manuscript and approved the final version of the manuscript.

## Acknowledgements

We gratefully acknowledge all the studies and databases that made GWAS summary data, expression data, and annotation data available as listed in the supplemental file. This research was conducted by using the UK Biobank resource under application numbers 29900.

