## Supplementary file for "Integrative Analyses Reveal Novel Disease-associated Loci and Genes for Idiopathic Pulmonary Fibrosis"

### Contents

**Supplementary Figures** ……………..……………..………………..………………..……..………..……... 3 - 12

Region mirror Manhattan plots for 14 local regions across 14 phenotypes with significant local genetic correlations with IPF tested using SUPERGNOVA (FDR < 0.05). P values from IPF GWAS summary statistics are plotted on the top and P values for the SNPs located within the significant SUPERGNOVA region is highlighted in blue. P values from another trait's summary statistics are plotted on the bottom and P values for the SNPs located within the significant SUPERGNOVA region is highlighted in yellow.

#### Supplementary Tables

S Table 1: Phenotype-level correlations ……………..……………..………………………………………… 13
S Table 2: Polygenic risk score correlations ……….…………..………..…………..…………..………... … 13
S Table 3: Global genetic correlations ……………..……………....……….……………………..…………… 14
S Table 4: Significant local regions from SUPERGNOVA ……………..……………..…………………..…..… …15
S Table 5: TWAS UTMOST results ……………..……………..……..……..…….....…….…………. …16
S Table 6: Differential gene expression analysis ……………..……………....…….……....…….…………..…. 17
S Table 7: Partitioned heritability results ……………..……………....…….……....…….…....………………18
S Table 8: *MAFK* and *SMAD2* target gene set from ChIP-Atlas ……………..……………......………………19
S Table 9A: Enrichment analysis of *MAFK* targets ……………..……………......……..……...……...……20 - 24
S Table 9B: Enrichment analysisof *SMAD2* targets ……………..……………......……..…......………..……25 - 28
S Table 10: GWAS study information ……………..……………......……..…....……….........……….……29

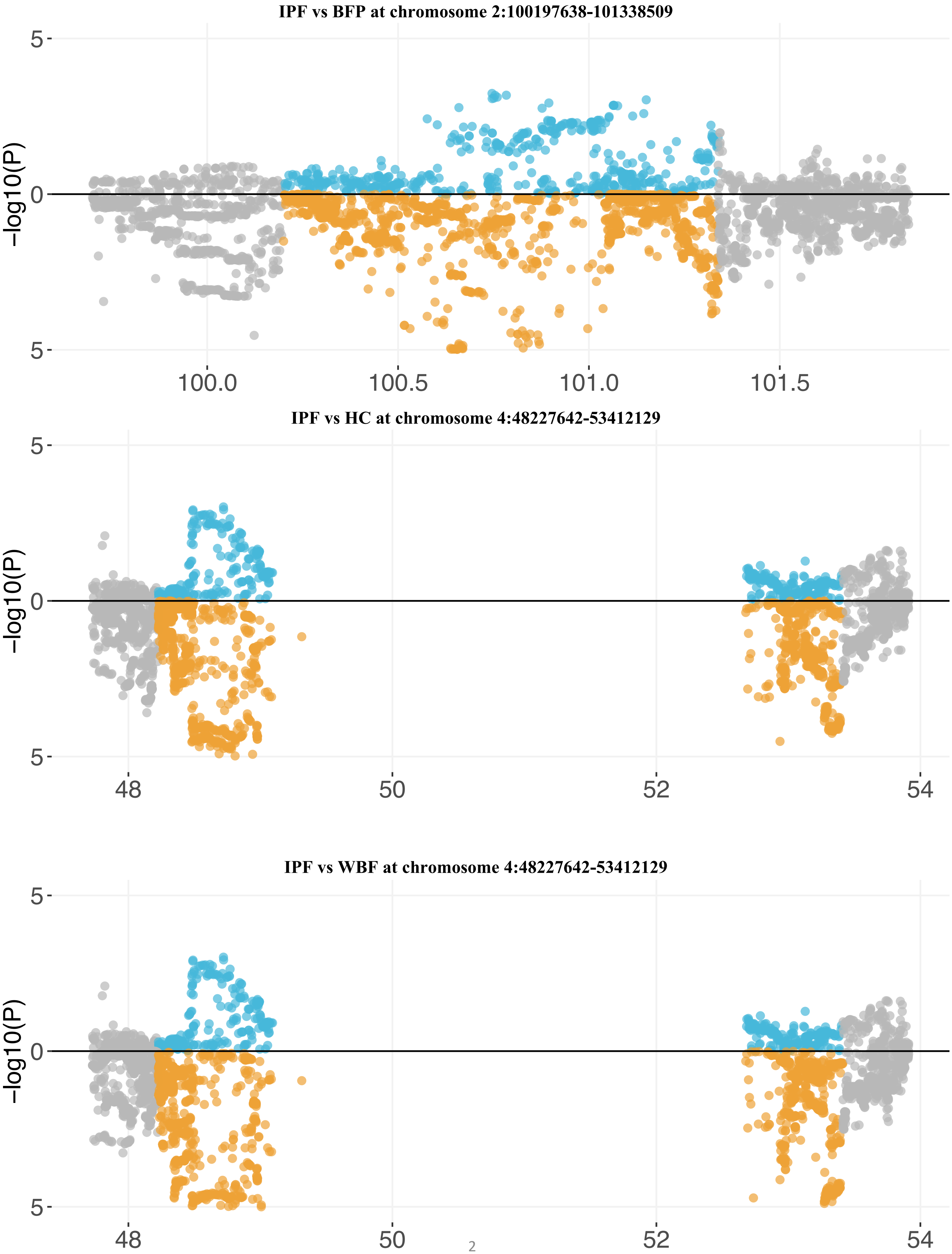
IPF vs BFP at chromosome 2:100197638-101338509
-log10(P)
5
0
5
100.0
100.5
101.0
101.5
IPF vs HC at chromosome 4:48227642-53412129
-log10(P)
5
0
5
48
50
52
54
IPF vs WBF at chromosome 4:48227642-53412129
-log10(P)
5
0
5
48
50
52
54

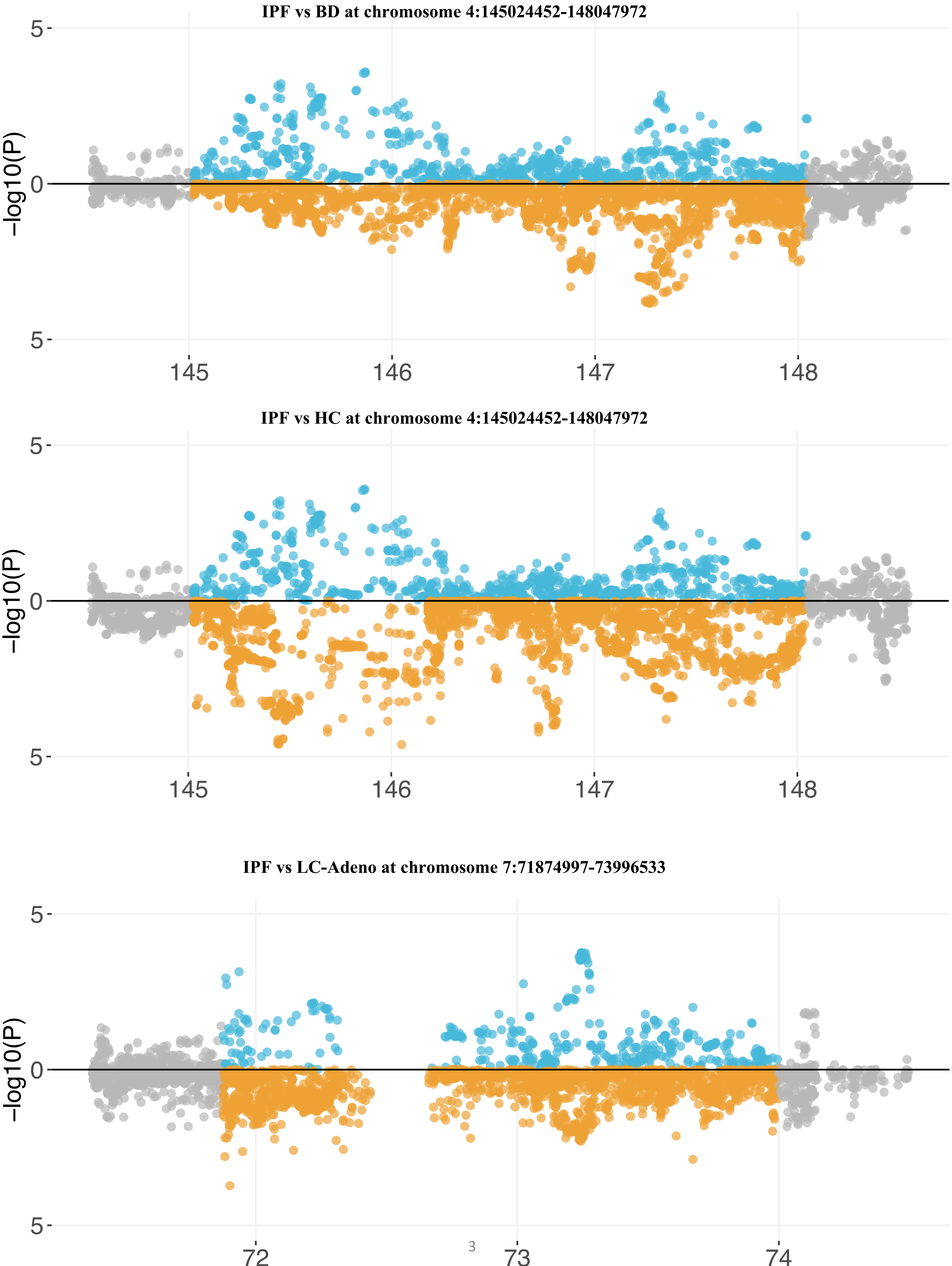
IPF vs BD at chromosome 4:145024452-148047972
-log10(P)
5
0
5
145
146
147
148
IPF vs HC at chromosome 4:145024452-148047972
-log10(P)
5
0
5
145
146
147
148
IPF vs LC-Adeno at chromosome 7:71874997-73996533
-log10(P)
5
0
5
72
73
74

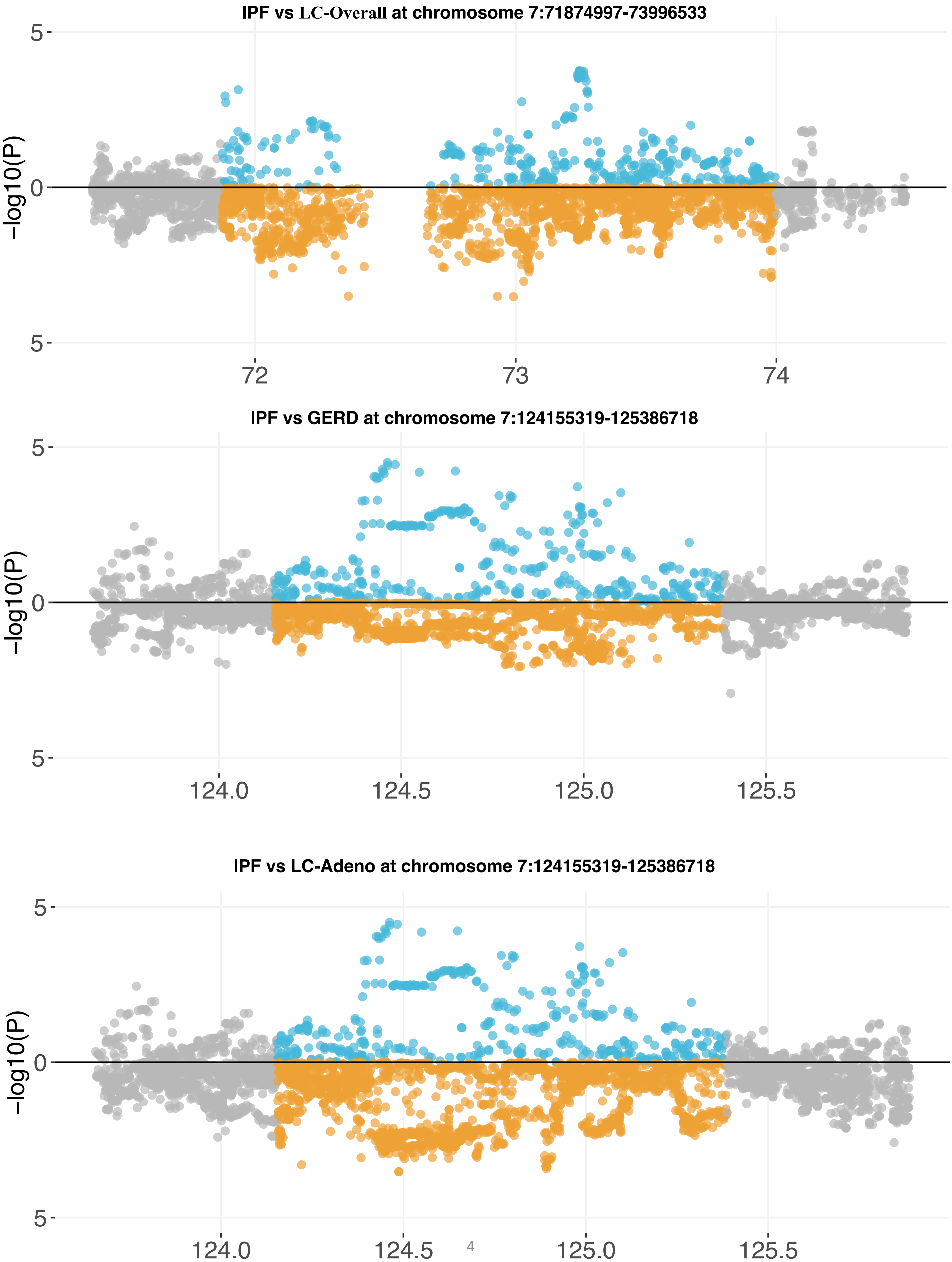

IPF vs LC-Overall at chromosome 7:71874997-73996533
5
0
5
-log10(P)
72
73
74
IPF vs GERD at chromosome 7:124155319-125386718
5
0
5
-log10(P)
124.0
124.5
125.0
125.5
IPF vs LC-Adeno at chromosome 7:124155319-125386718
5
0
5
-log10(P)
124.0
124.5
125.0
125.5

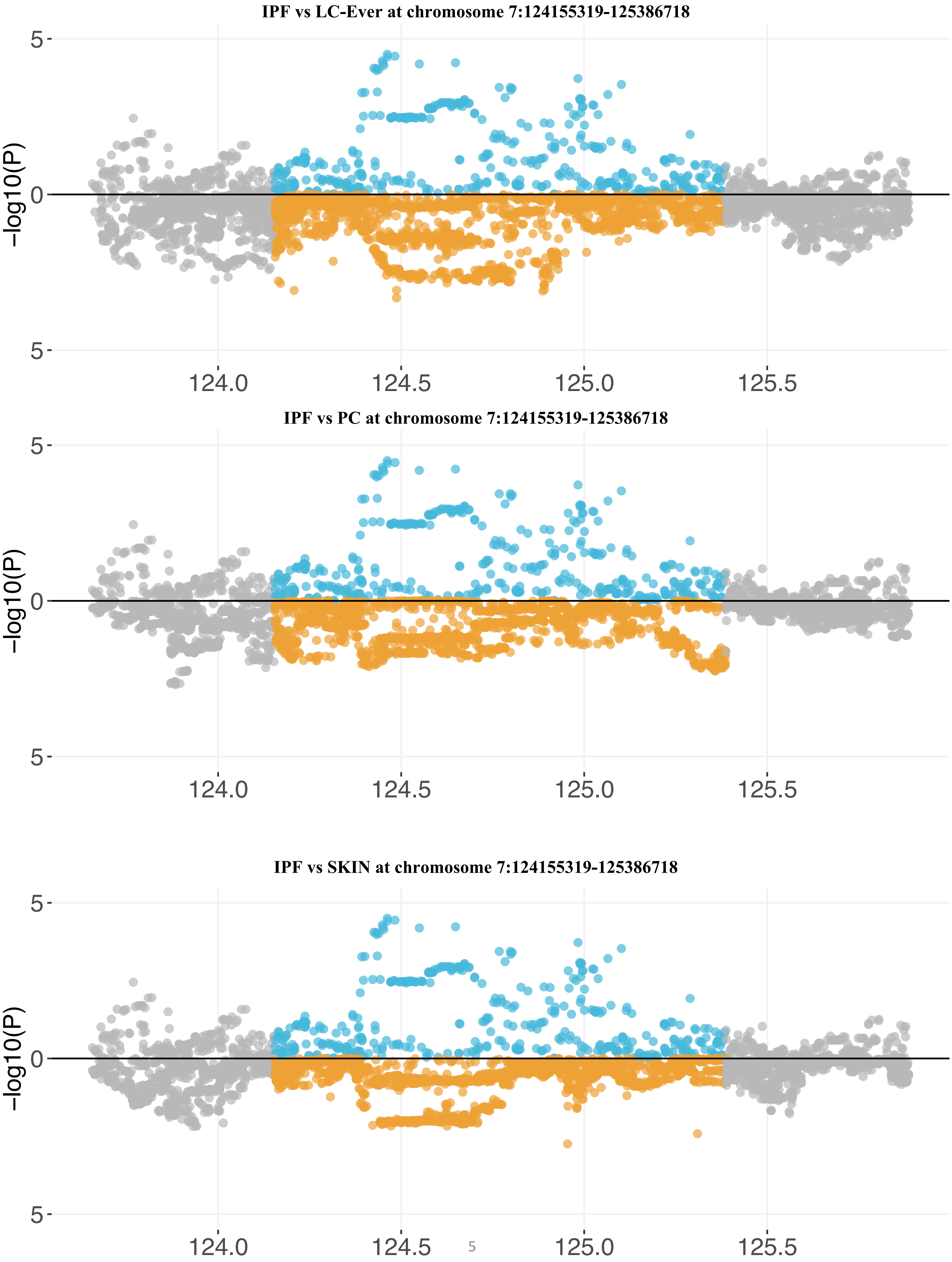
IPF vs LC-Ever at chromosome 7:124155319-125386718
IPF vs PC at chromosome 7:124155319-125386718
IPF vs SKIN at chromosome 7:124155319-125386718
−log10(P)
5
0
5
124.0
124.5
125.0
125.5

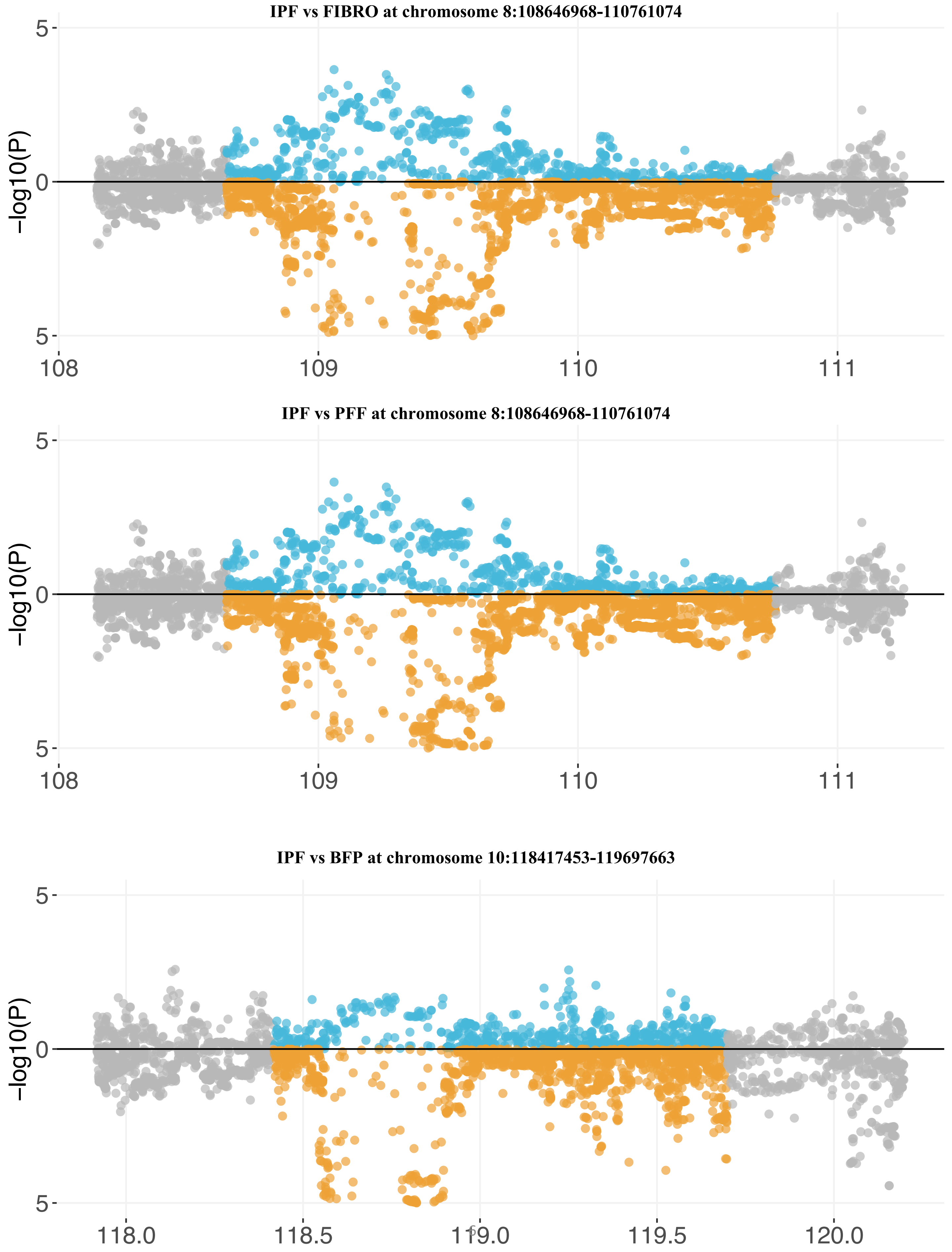
IPF vs FIBRO at chromosome 8:108646968-110761074
-log10(P)
5
0
5
108
109
110
111
IPF vs PFF at chromosome 8:108646968-110761074
-log10(P)
5
0
5
108
109
110
111
IPF vs BFP at chromosome 10:118417453-119697663
-log10(P)
5
0
5
118.0
118.5
119.0
119.5
120.0

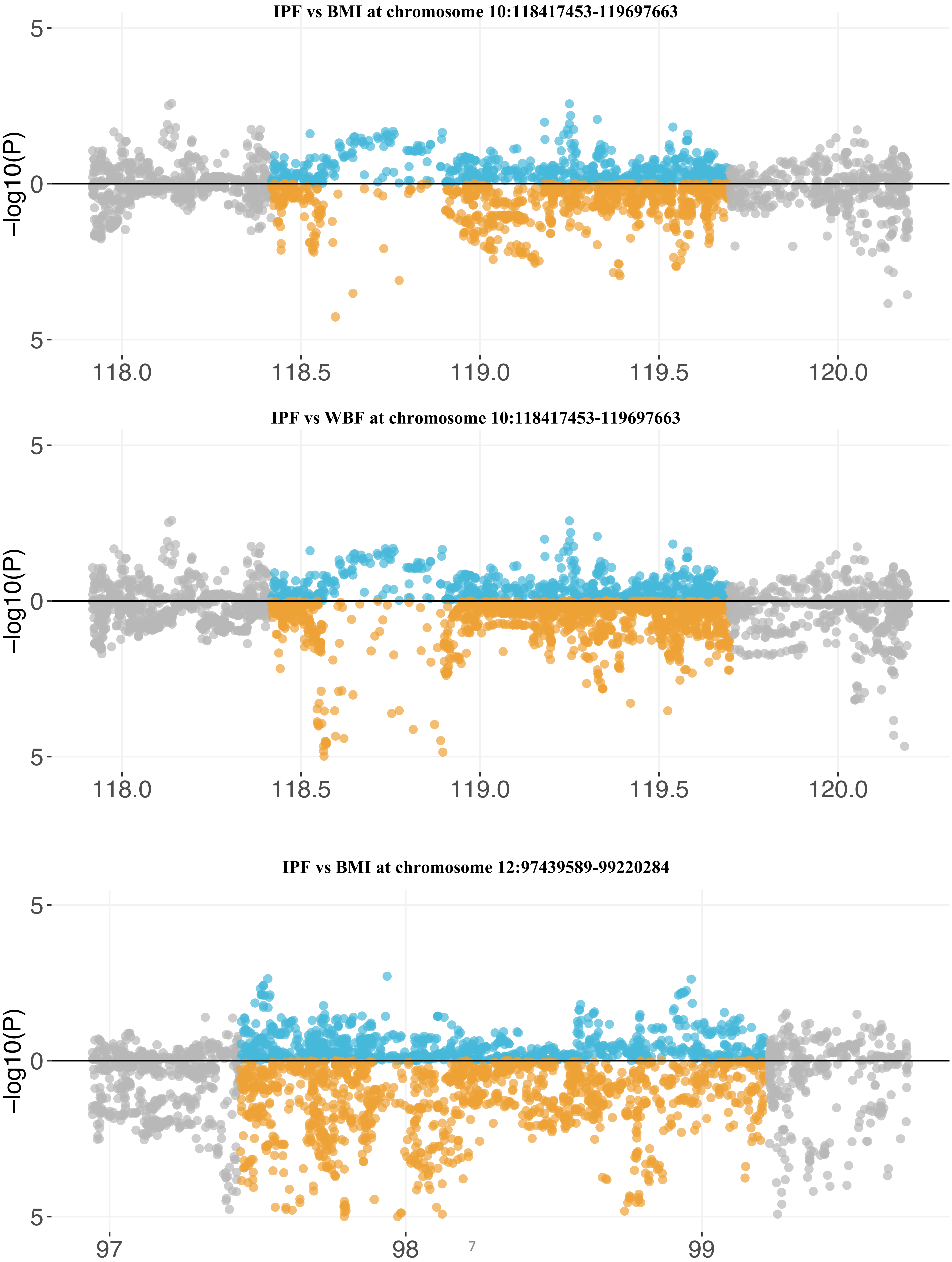
IPF vs BMI at chromosome 10:118417453-119697663
IPF vs WBF at chromosome 10:118417453-119697663
IPF vs BMI at chromosome 12:97439589-99220284
−log10(P)
5
0
5
118.0
118.5
119.0
119.5
120.0
97
98
99

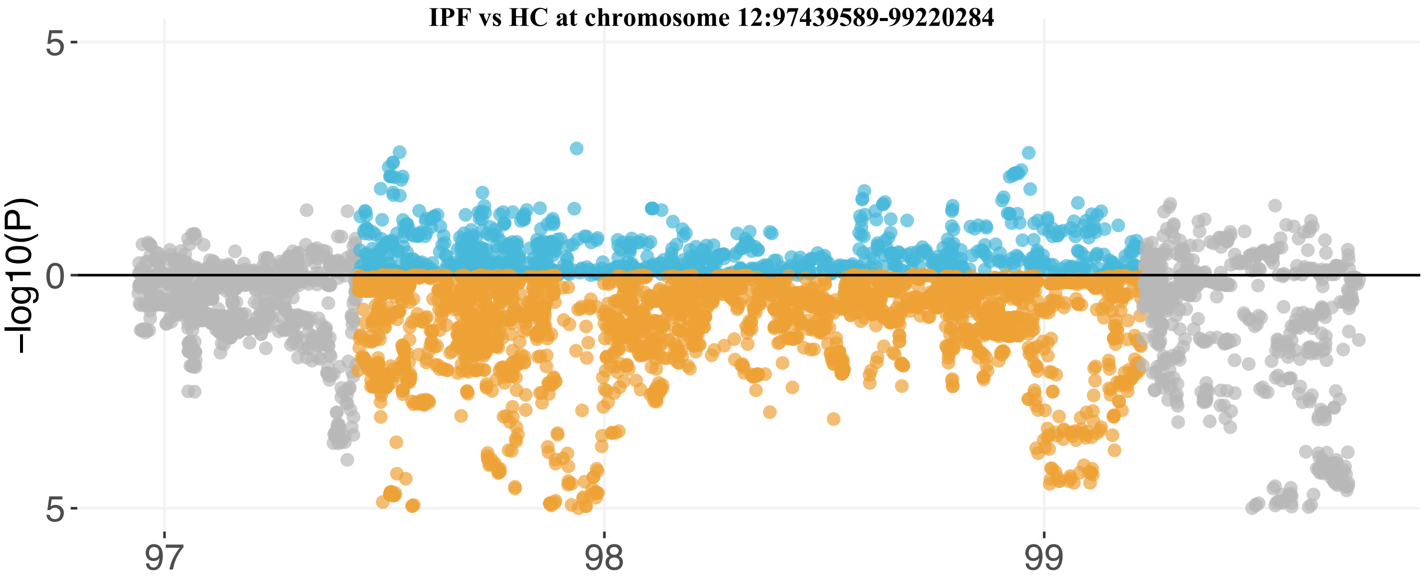
IPF vs HC at chromosome 12:97439589-99220284
-log10(P)
5
0
5
97
98
99

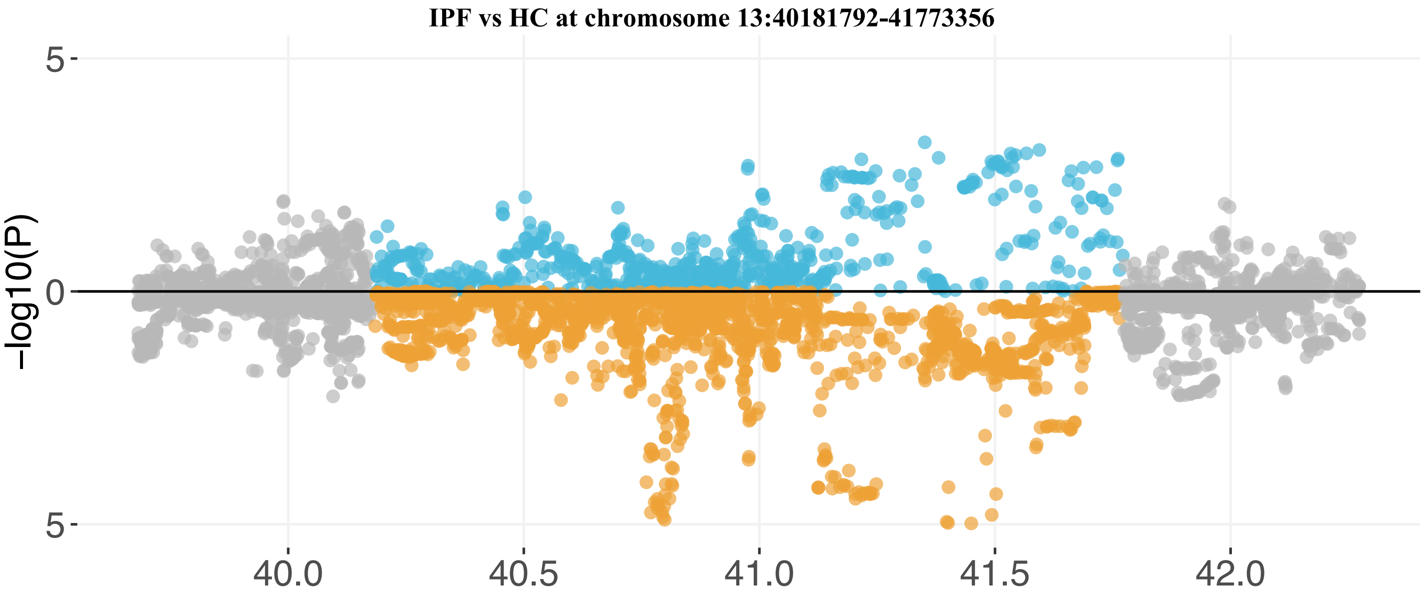
IPF vs HC at chromosome 13:40181792-41773356
-log10(P)
5
0
5
40.0
40.5
41.0
41.5
42.0

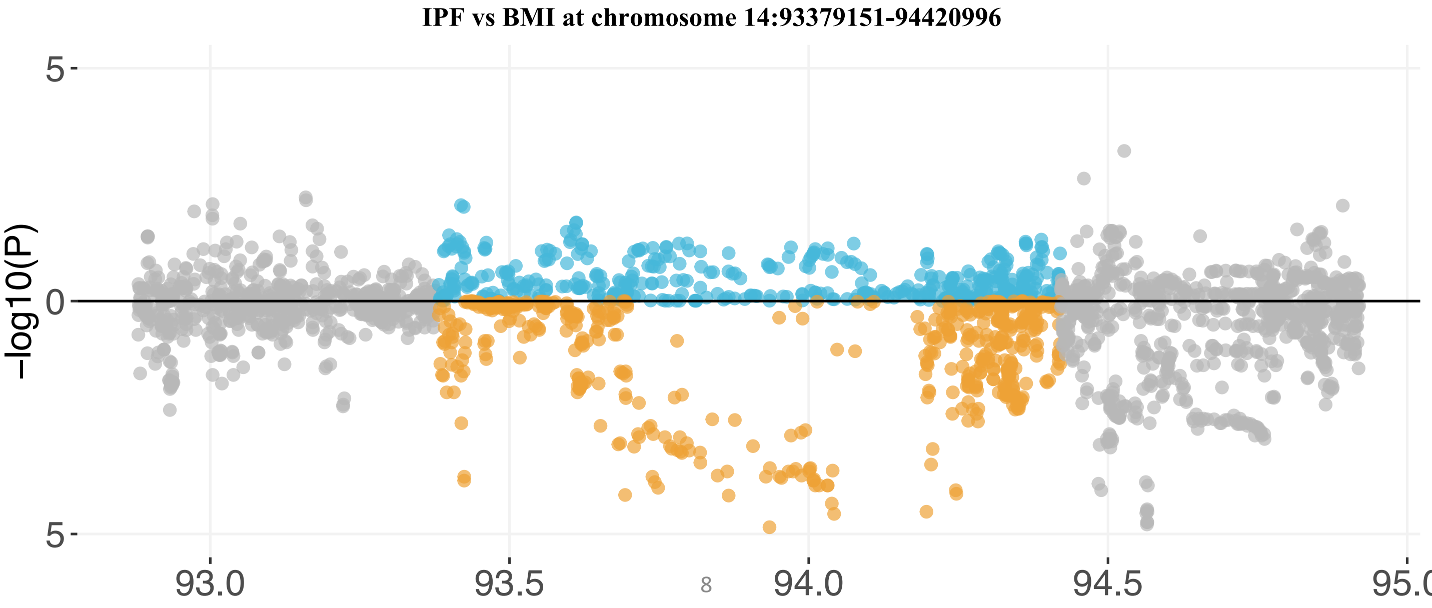
IPF vs BMI at chromosome 14:93379151-94420996
-log10(P)
5
0
5
93.0
93.5
94.0
94.5
95.0

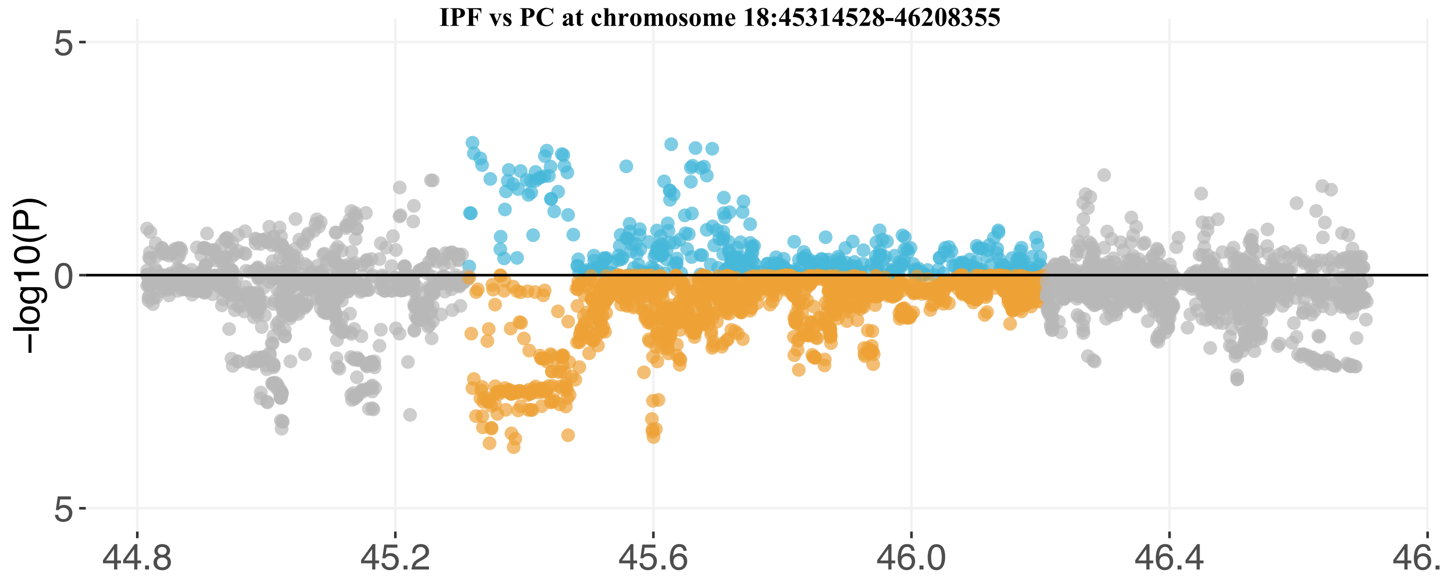
IPF vs PC at chromosome 18:45314528-46208355
5
0
5
−log10(P)
44.8
45.2
45.6
46.0
46.4
46.

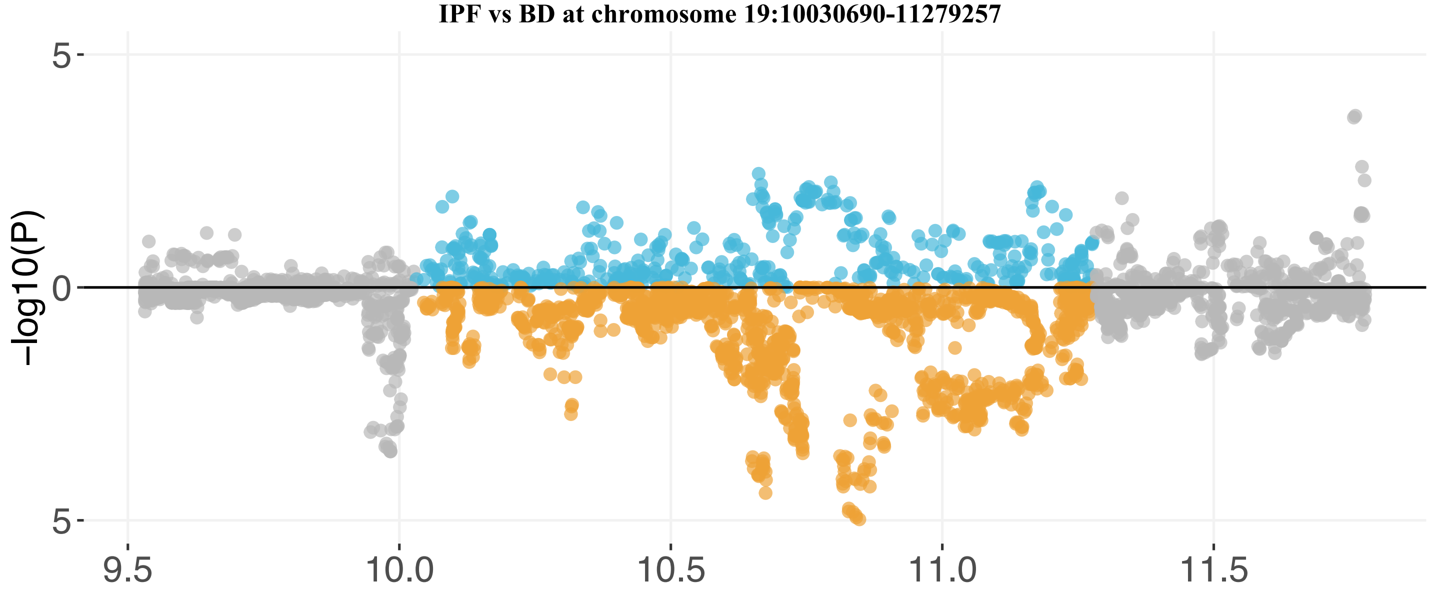
IPF vs BD at chromosome 19:10030690-11279257
5
0
5
−log10(P)
9.5
10.0
10.5
11.0
11.5

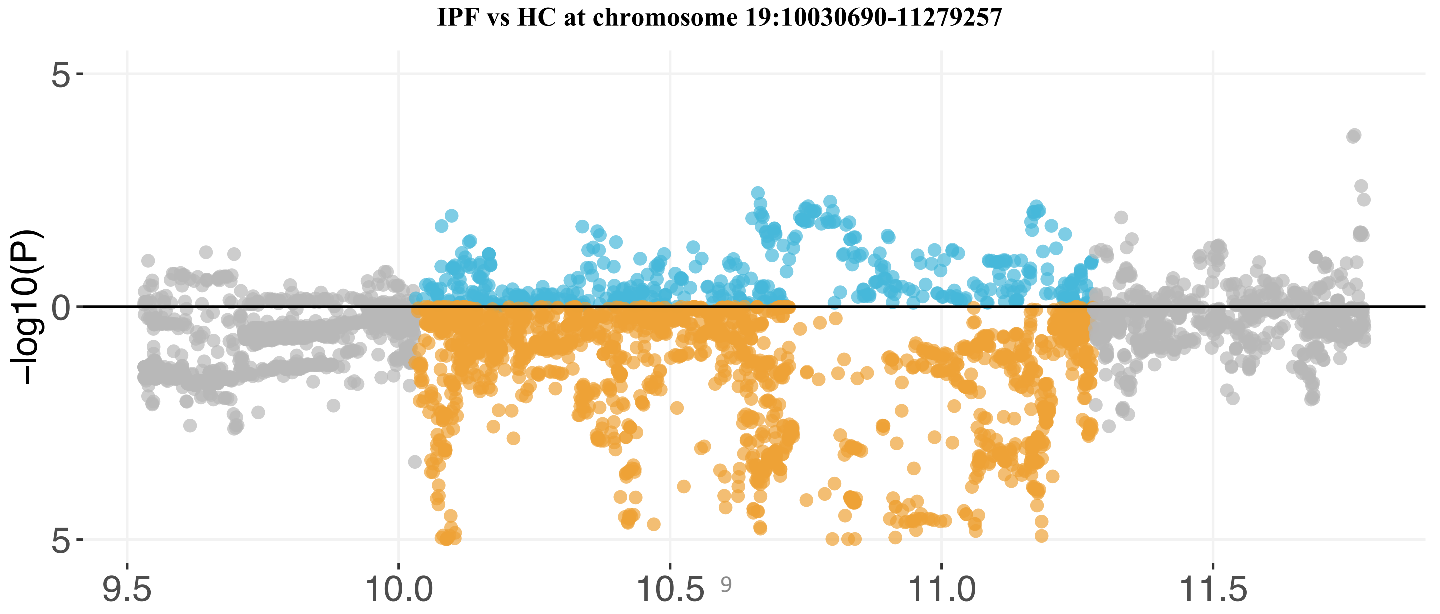
IPF vs HC at chromosome 19:10030690-11279257
5
0
5
−log10(P)
9.5
10.0
10.5
11.0
11.5

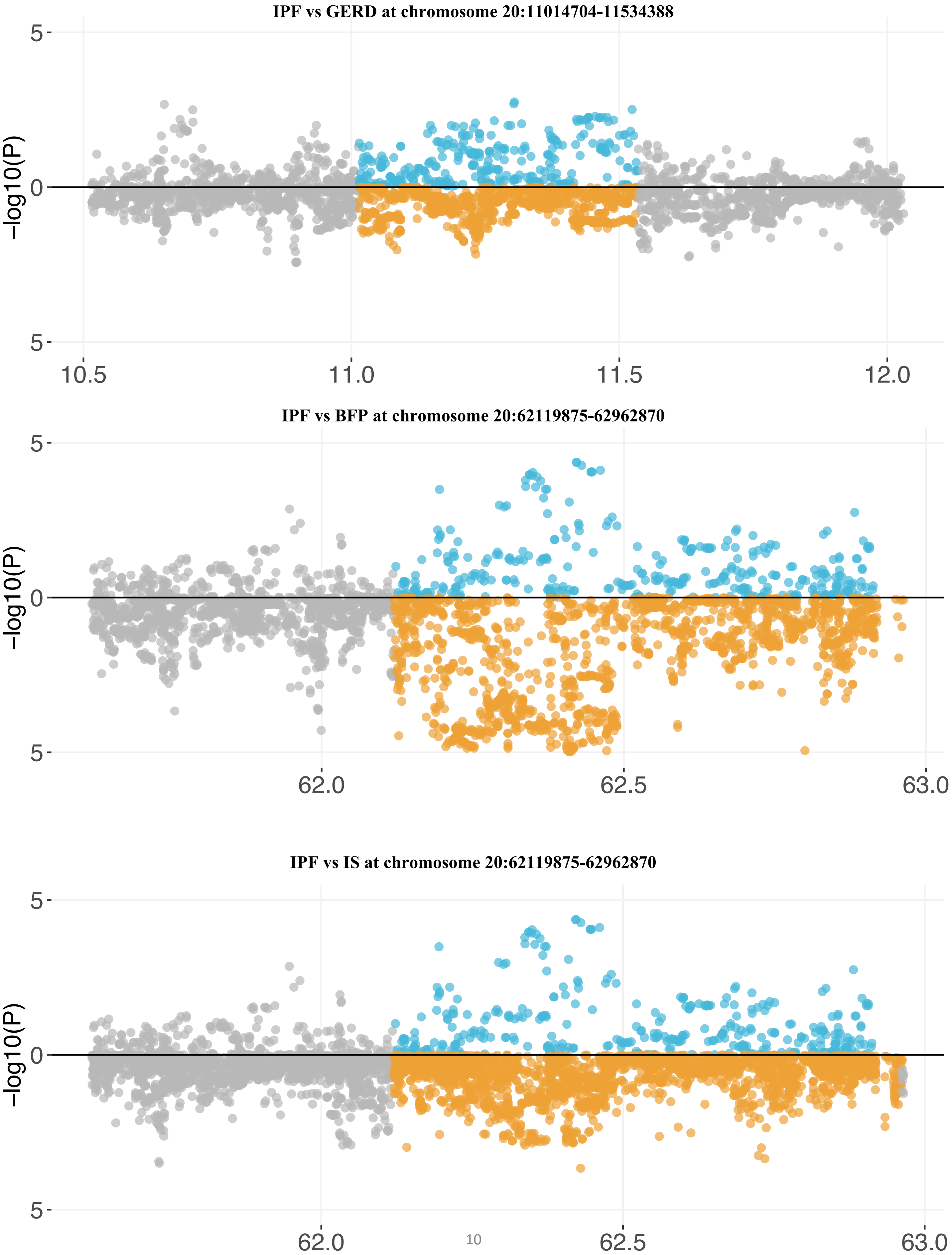

IPF vs GERD at chromosome 20:11014704-11534388
-log10(P)
5
0
5
10.5
11.0
11.5
12.0
IPF vs BFP at chromosome 20:62119875-62962870
-log10(P)
5
0
5
62.0
62.5
63.0
IPF vs IS at chromosome 20:62119875-62962870
-log10(P)
5
0
5
62.0
62.5
63.0

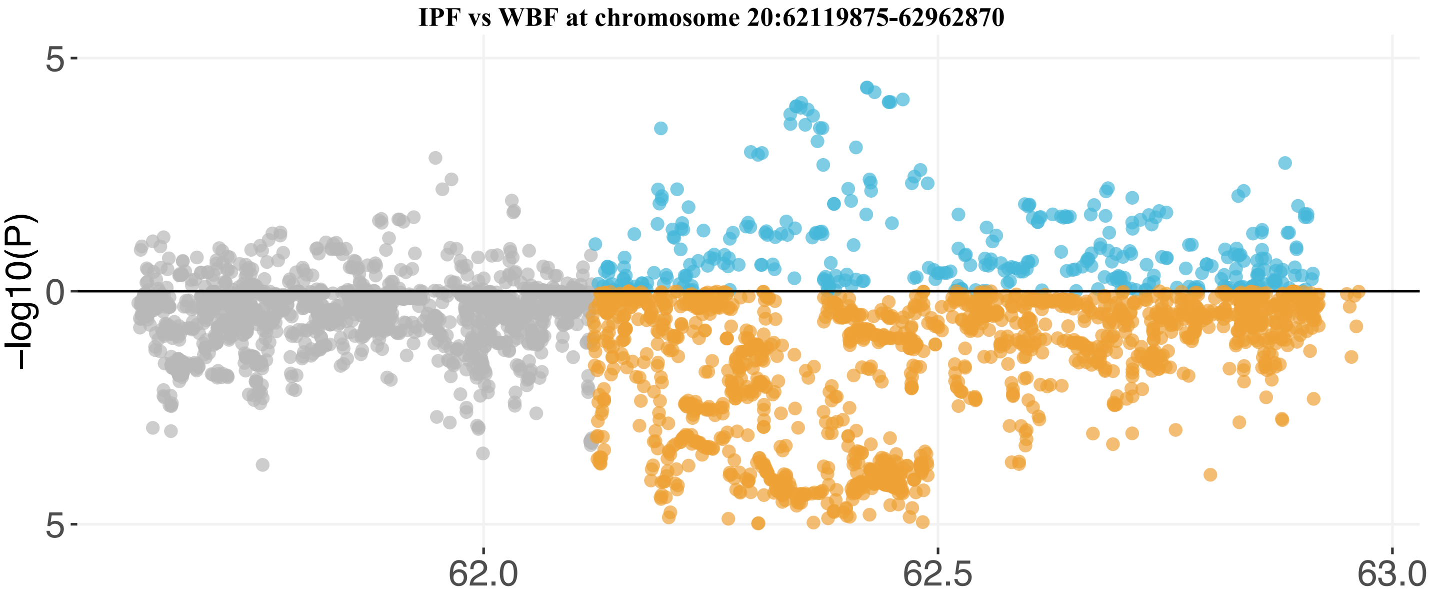

IPF vs WBF at chromosome 20:62119875-62962870
5
0
5
-log10(P)
62.0
62.5
63.0

#### S Table 1: Phenotype-level correlations between UK Biobank traits and IPF (FDR < 0.05)

| Phenotype | R square | P value | Coefficient | Adjusted P value (FDR) | Adjusted P value (Bonferroni) |
|---|---|---|---|---|---|
| Doctor diagnosed fibrosing alveolitis/unspecified alveolitis | 0.046 | 4.66E-36 | 5.57 | 3.12E-33 | 3.12E-33 |
| Systemic Lupus Erythematosus | 0.011 | 3.40E-18 | 4.46 | 1.14E-15 | 2.27E-15 |
| Doctor diagnosed emphysema | 0.025 | 8.23E-18 | 3.44 | 1.79E-15 | 5.51E-15 |
| Doctor diagnosed bronchiectasis | 0.029 | 1.07E-17 | 2.78 | 1.79E-15 | 7.16E-15 |
| Doctor diagnosed COPD | 0.029 | 1.75E-16 | 2.49 | 2.34E-14 | 1.17E-13 |
| Number of treatments/medications taken | 0.025 | 4.72E-14 | 0.18 | 5.26E-12 | 3.16E-11 |
| Doctor diagnosed tuberculosis | 0.015 | 2.43E-10 | 2.70 | 2.32E-08 | 1.63E-07 |
| Age at recruitment | 0.033 | 3.29E-10 | 0.12 | 2.75E-08 | 2.20E-07 |
| Year of birth | 0.032 | 4.34E-10 | -0.11 | 3.23E-08 | 2.90E-07 |
| Doctor diagnosed sarcoidosis | 0.013 | 7.38E-09 | 2.68 | 4.94E-07 | 4.94E-06 |
| Nap during day | 0.017 | 6.41E-08 | 0.85 | 3.65E-06 | 4.29E-05 |
| Lymphocyte percentage | 0.020 | 6.56E-08 | 0.00 | 3.65E-06 | 4.39E-05 |
| Number of self-reported non-cancer illnesses | 0.013 | 7.10E-08 | 0.20 | 3.65E-06 | 4.75E-05 |
| Albumin | 0.012 | 1.27E-07 | -0.17 | 6.07E-06 | 8.50E-05 |
| Medication for cholesterol, blood pressure or diabetes | 0.032 | 2.19E-07 | 0.44 | 9.77E-06 | 0.00014651 |
| Long-standing illness, disability or infirmity | 0.017 | 4.57E-07 | 1.07 | 1.91E-05 | 0.00030573 |
| Taking other prescription medications | 0.018 | 7.43E-07 | 1.22 | 2.92E-05 | 0.00049707 |
| C_reactive_protein | 0.006 | 8.55E-07 | 0.04 | 3.18E-05 | 0.000572 |
| Doctor diagnosed cystic fibrosis | 0.008 | 2.12E-06 | 6.73 | 7.46E-05 | 0.00141828 |
| Red blood cell (erythrocyte) distribution width | 0.010 | 3.66E-06 | 0.00 | 0.00012 | 0.00244854 |
| Other serious medical condition/disability diagnosed by doctor | 0.013 | 4.20E-06 | 0.99 | 0.00013 | 0.0028098 |
| Neutrophill count | 0.011 | 7.36E-06 | 0.00 | 0.00022 | 0.00492384 |
| Cystatin_C | 0.004 | 1.49E-05 | 0.74 | 0.00043 | 0.0099681 |
| Chest pain or discomfort | 0.010 | 1.81E-05 | 0.97 | 0.00050 | 0.0121089 |
| Overall health rating | 0.010 | 4.88E-05 | 0.55 | 0.0013 | 0.0326472 |
| Total Cholesterol | 0.006 | 1.00E-04 | -0.31 | 0.0026 | 0.0669 |
| White blood cell (leukocyte) count | 0.008 | 0.00016 | 0.00 | 0.0038 | 0.10382679 |
| Monocyte count | 0.005 | 0.00017 | 0.01 | 0.0041 | 0.11571626 |
| COPD | 0.005 | 0.00020 | 0.75 | 0.0047 | 0.13556081 |
| Waist circumference | 0.008 | 0.00029 | 0.03 | 0.0064 | 0.19288675 |
| LC | 0.003 | 0.00037 | 1.81 | 0.0079 | 0.24563071 |
| Eosinophill count | 0.006 | 0.00045 | 0.02 | 0.0094 | 0.30113965 |
| Forced vital capacity (FVC) | 0.009 | 0.00057 | -0.44 | 0.011 | 0.37820912 |
| LDL | 0.005 | 0.00064 | -0.36 | 0.013 | 0.42796131 |
| Forced vital capacity (FVC), Best measure | 0.011 | 0.00077 | 0.00 | 0.015 | 0.51550531 |
| Forced expiratory volume in 1-second (FEV1) | 0.009 | 0.00091 | -0.53 | 0.017 | 0.60714292 |
| Diabetes diagnosed by doctor | 0.005 | 0.00092 | 1.06 | 0.017 | 0.61433467 |
| Surgery on leg arteries (other than for varicose veins) | 0.034 | 0.0010 | -2.91 | 0.018 | 0.69112182 |
| Average monthly intake of other alcoholic drinks | 0.033 | 0.0013 | 0.38 | 0.021 | 0.84787655 |
| Doctor diagnosed alpha-1 antitrypsin deficiency | 0.003 | 0.0013 | 3.29 | 0.021 | 0.854503 |
| Creatinine | 0.002 | 0.0014 | 0.00 | 0.023 | 0.92877337 |
| Forced expiratory volume in 1-second (FEV1), Best measure | 0.009 | 0.0017 | -0.01 | 0.027 | 1 |
| Eye problems/disorders | 0.013 | 0.0017 | 0.16 | 0.027 | 1 |
| IGF_1 | 0.004 | 0.0024 | -0.05 | 0.036 | 1 |
| Neutrophill percentage | 0.007 | 0.0026 | 0.00 | 0.039 | 1 |
| Other eye problems | 0.005 | 0.0034 | 0.72 | 0.049 | 1 |

#### S Table 2: Correlations between UK Biobank traits and IPF polygenic risk score (FDR < 0.05)

| Phenotype | R square | P value | Coefficient | Adjusted P value (FDR) | Adjusted P value (Bonferroni) |
|---|---|---|---|---|---|
| Monocyte percentage | -5.59E-06 | 7.46E-09 | 1.69E-10 | 4.99E-06 | 4.99E-06 |
| Monocyte count | -3.11E-06 | 1.63E-05 | 1.51E-09 | 0.00545235 | 0.0109047 |
| Mean corpuscular haemoglobin concentration | -2.57E-06 | 8.75E-05 | -2.72E-10 | 0.0195125 | 0.0585375 |

**S Table 3: Global genetic correlations between UK Biobank traits and IPF (FDR < 0.05)**

| Trait | $h^2$ | Rho corrected | Corr corrected | Pvalue corrected | p_fdr |
|---|---|---|---|---|---|
| Fibroblastic disorders | 0.017 | 0.027 | 0.42 | 1.50E-06 | 0.001 |
| Ischaemic Stroke | 0.062 | 0.051 | 0.41 | 1.09E-05 | 0.002 |
| Palmar fascial fibromatosis [Dupuytren] | 0.017 | 0.024 | 0.36 | 1.80E-05 | 0.002 |
| Other/unspecified dorsalgia | 0.004 | -0.016 | -0.51 | 4.72E-05 | 0.004 |
| Body Mass Index | 0.164 | 0.024 | 0.17 | 6.14E-05 | 0.004 |
| Arm fat percentage | 0.194 | 0.030 | 0.13 | 6.20E-05 | 0.004 |
| Diagnoses - main ICD10: S42 Fracture of shoulder and upper arm | 0.002 | 0.014 | 0.66 | 7.70E-05 | 0.004 |
| Insomnia | 0.042 | -0.026 | -0.25 | 0.00021 | 0.010 |
| Type 2 diabetes with ophthalmic complications | 0.001 | 0.013 | 0.69 | 0.00025 | 0.010 |
| Body fat percentage | 0.200 | 0.036 | 0.16 | 0.00026 | 0.010 |
| Whole body fat mass | 0.207 | 0.033 | 0.14 | 0.00041 | 0.014 |
| Total protein (quantile) | 0.190 | 0.006 | 0.03 | 0.00044 | 0.014 |
| Hip circumference | 0.193 | 0.036 | 0.16 | 0.00065 | 0.019 |
| Diagnoses - main ICD10: E04 Other non-toxic goitre | 0.005 | -0.013 | -0.34 | 0.00075 | 0.019 |
| Diseases of the nervous system | 0.014 | 0.013 | 0.21 | 0.00078 | 0.019 |
| Malignant neoplasm of respiratory system and intrathoracic organs | 0.003 | 0.019 | 0.72 | 0.00085 | 0.019 |
| Diagnoses - main ICD10: C34 Malignant neoplasm of bronchus and lung | 0.003 | 0.014 | 0.53 | 0.00086 | 0.019 |
| Malignant melanoma of skin | 0.004 | 0.016 | 0.50 | 0.00089 | 0.019 |
| Diagnoses - main ICD10: K52 Other non-infective gastro-enteritis and colitis | 0.006 | 0.012 | 0.32 | 0.00113 | 0.023 |
| Creatinine (quantile) | 0.187 | -0.005 | -0.02 | 0.00165 | 0.030 |
| SHBG (quantile) | 0.185 | -0.005 | -0.02 | 0.00169 | 0.030 |
| Diagnoses - main ICD10: C61 Malignant neoplasm of prostate | 0.010 | -0.014 | -0.27 | 0.00175 | 0.030 |
| Non-cancer illness code, self-reported: prolapsed disc/slipped disc | 0.007 | 0.013 | 0.30 | 0.00182 | 0.030 |
| Non-cancer illness code, self-reported: rheumatoid arthritis | 0.012 | -0.014 | -0.26 | 0.00257 | 0.039 |
| Hernia | 0.017 | -0.019 | -0.29 | 0.00264 | 0.039 |
| Asthma-related pneumonia | 0.003 | 0.012 | 0.40 | 0.00344 | 0.049 |

**S Table 4: Significant local regions from SUPERGNOVA (FDR < 0.05)**

| GRCh37 Chr | GRCh37 Start | GRCh37 End | Rho | Correlation | IPF $h^2$ | Trait $h^2$ | Variance | P value | Number of SNP | Trait | Adjusted p value |
|---|---|---|---|---|---|---|---|---|---|---|---|
| 2 | 100197638 | 101338509 | 0.0009181 | 1.076084853 | 0.0022 | 0.00033 | 5.37E-08 | 7.41E-05 | 450 | BFP | 0.040 |
| 4 | 48227642 | 53412129 | -0.0004397 | -1.148381067 | 0.0007 | 0.00020 | 1.29E-08 | 1.07E-04 | 398 | HC | 0.048 |
| 4 | 48227642 | 53412129 | -0.0004624 | -1.171372833 | 0.0007 | 0.00022 | 1.37E-08 | 7.64E-05 | 398 | WBF | 0.040 |
| 4 | 145024452 | 148047972 | 0.00152152 | 1.319656624 | 0.0027 | 0.00049 | 1.36E-07 | 3.64E-05 | 800 | BD | 0.027 |
| 4 | 145024452 | 148047972 | 0.00153375 | 0.893286257 | 0.0030 | 0.00099 | 1.11E-07 | 4.35E-06 | 822 | HC | 0.007 |
| 7 | 71874997 | 73996533 | -0.000992 | -1.390732517 | 0.0034 | 0.00015 | 5.61E-08 | 2.83E-05 | 388 | LC-Adeno | 0.026 |
| 7 | 71874997 | 73996533 | -0.0012313 | -1.020456228 | 0.0032 | 0.00045 | 8.41E-08 | 2.19E-05 | 358 | LC-Overall | 0.025 |
| 7 | 124155319 | 125386718 | 0.00031845 | 1.622580856 | 0.0027 | 0.00001 | 5.96E-09 | 3.71E-05 | 376 | GERD | 0.027 |
| 7 | 124155319 | 125386718 | -0.0010126 | -0.9591298 | 0.0028 | 0.00040 | 3.55E-08 | 7.66E-08 | 376 | LC-Adeno | 0.000 |
| 7 | 124155319 | 125386718 | -0.0011691 | -1.0546343 | 0.0028 | 0.00044 | 7.35E-08 | 1.62E-05 | 376 | LC-Ever | 0.018 |
| 7 | 124155319 | 125386718 | -0.0003354 | -1.264624298 | 0.0027 | 0.00003 | 5.57E-09 | 7.04E-06 | 376 | PC | 0.010 |
| 7 | 124155319 | 125386718 | -0.000337 | -1.2474198 | 0.0027 | 0.00003 | 4.69E-09 | 8.73E-07 | 376 | SKIN | 0.003 |
| 8 | 108646968 | 110761074 | 0.00140409 | 1.095712503 | 0.0021 | 0.00077 | 6.49E-08 | 3.55E-08 | 722 | FIBRO | 0.000 |
| 8 | 108646968 | 110761074 | 0.00138822 | 1.09978966 | 0.0021 | 0.00074 | 6.24E-08 | 2.77E-08 | 722 | PFF | 0.000 |
| 10 | 118417453 | 119697663 | 0.00076178 | 3.466637912 | 0.0001 | 0.00040 | 3.44E-08 | 4.04E-05 | 596 | BFP | 0.028 |
| 10 | 118417453 | 119697663 | 0.00063527 | 4.373532223 | 0.0001 | 0.00029 | 2.12E-08 | 1.25E-05 | 521 | BMI | 0.016 |
| 10 | 118417453 | 119697663 | 0.0008632 | 3.646813319 | 0.0001 | 0.00046 | 3.41E-08 | 2.93E-06 | 596 | WBF | 0.006 |
| 12 | 97439589 | 99220284 | -0.0007808 | -1.618998426 | 0.0006 | 0.00036 | 3.71E-08 | 5.02E-05 | 801 | BMI | 0.032 |
| 12 | 97439589 | 99220284 | -0.0009282 | -2.074365504 | 0.0004 | 0.00046 | 4.93E-08 | 2.90E-05 | 873 | HC | 0.026 |
| 13 | 40181792 | 41773356 | 0.00071865 | 0.754733444 | 0.0037 | 0.00025 | 3.24E-08 | 6.52E-05 | 629 | HC | 0.039 |
| 14 | 93379151 | 94420996 | 0.00054221 | 1.748899691 | 0.0002 | 0.00050 | 1.93E-08 | 9.68E-05 | 397 | BMI | 0.045 |
| 18 | 45314528 | 46208355 | 0.00043225 | 1.446763395 | 0.0014 | 0.00006 | 1.20E-08 | 8.24E-05 | 410 | PC | 0.042 |
| 19 | 10030690 | 11279257 | -0.0018445 | -1.247563343 | 0.0015 | 0.00141 | 1.53E-07 | 2.40E-06 | 361 | BD | 0.006 |
| 19 | 10030690 | 11279257 | 0.00105941 | 1.132095119 | 0.0016 | 0.00053 | 5.17E-08 | 3.17E-06 | 423 | HC | 0.006 |
| 20 | 11014704 | 11534388 | -0.0003089 | -1.68816263 | 0.0019 | 0.00002 | 6.20E-09 | 8.78E-05 | 288 | GERD | 0.043 |
| 20 | 62119875 | 62962870 | 0.00137656 | 0.893169382 | 0.0054 | 0.00044 | 1.09E-07 | 2.94E-05 | 322 | BFP | 0.026 |
| 20 | 62119875 | 62962870 | 0.00225362 | 1.068203883 | 0.0054 | 0.00083 | 2.94E-07 | 3.28E-05 | 324 | IS | 0.027 |
| 20 | 62119875 | 62962870 | 0.00114659 | 0.871566982 | 0.0054 | 0.00032 | 8.26E-08 | 6.64E-05 | 322 | WBF | 0.039 |

#### S Table 5: TWAS UTMOST significant genes from joint analysis of 44 tissues from GTEx

| Gene | Test score | p value | GRCh37 chromosome | GRCh37 position | Adjusted p value | Identified GWAS hit | Identified DEG |
|---|---|---|---|---|---|---|---|
| AKAP13 | 18.22 | 7.94E-09 | 15 | 85923802-86292586 | 3.10E-03 | T | T |
| ANO9 | 31.33 | 7.11E-15 | 11 | 417930-442011 | 2.78E-09 | | T |
| ARHGAP27 | 25.90 | 2.15E-12 | 17 | 43471268-43511787 | 8.38E-07 | T | T |
| ARL17A | 16.71 | 1.18E-08 | 17 | 43697710-43913194 | 4.62E-03 | | |
| B4GALNT4 | 15.85 | 4.58E-08 | 11 | 369795-382117 | 1.79E-02 | | T |
| BAHD1 | 26.24 | 2.26E-12 | 15 | 40731920-40760441 | 8.84E-07 | | |
| BMF | 133.41 | 1.83E-11 | 15 | 40380091-40401093 | 7.14E-06 | | |
| BRSK2 | 125.77 | 4.15E-10 | 11 | 1411129-1483919 | 1.62E-04 | T | |
| CDHR5 | 28.26 | 4.36E-13 | 11 | 616565-626078 | 1.70E-07 | | |
| CNPY4 | 14.97 | 1.01E-07 | 7 | 99717236-99723134 | 3.96E-02 | | T |
| CRHR1 | 23.95 | 2.71E-11 | 17 | 43697710-43913194 | 1.06E-05 | T | |
| CTSD | 106.87 | 4.09E-11 | 11 | 1773982-1785222 | 1.60E-05 | | T |
| DEPTOR | 17.37 | 1.03E-08 | 8 | 120885900-121063157 | 4.04E-03 | T | |
| EFCAB4A | 19.81 | 8.39E-10 | 11 | 826144-831991 | 3.28E-04 | | T |
| FAM13A | 21.22 | 2.84E-10 | 4 | 89647106-90032549 | 1.11E-04 | T | T |
| GJC1 | 23.99 | 1.09E-11 | 17 | 42875816-42908184 | 4.24E-06 | | |
| HRAS | 15.22 | 6.70E-08 | 11 | 532242-537287 | 2.62E-02 | | |
| IRF7 | 120.20 | 2.22E-11 | 11 | 612553-615999 | 8.67E-06 | | T |
| KRTAP5-1 | 77.98 | 2.50E-11 | 11 | 1605572-1606513 | 9.77E-06 | | T |
| LMNTD2 | 15.87 | 7.04E-08 | 11 | 554850-560779 | 2.75E-02 | | |
| LRRC37A | 667.77 | 2.27E-11 | 17 | 44370099-44415160 | 8.85E-06 | T | |
| MAFK | 15.58 | 5.12E-08 | 7 | 1570350-1582679 | 2.00E-02 | | |
| MAPT | 305.78 | 2.06E-11 | 17 | 43971748-44105700 | 8.04E-06 | T | |
| MYNN | 507.64 | 2.34E-11 | 3 | 169490619-169507504 | 9.13E-06 | | T |
| PAK6 | 161.50 | 3.45E-08 | 4 | 40509629-40569688 | 1.35E-02 | | |
| PKD2 | 17.99 | 8.56E-09 | 4 | 88928799-88998931 | 3.34E-03 | | T |
| PLEKHM1 | 340.62 | 2.36E-11 | 17 | 43513266-43568146 | 9.23E-06 | | T |
| PPM1K | 16.49 | 5.33E-08 | 4 | 89178772-89205921 | 2.08E-02 | | T |
| PRR33 | 488.77 | 1.98E-11 | 11 | 1910375-1912084 | 7.72E-06 | | |
| SCT | 83.37 | 1.09E-11 | 11 | 626313-627173 | 4.26E-06 | | |
| SEMA6B | 18.90 | 6.27E-09 | 19 | 4542600-4559820 | 2.45E-03 | | T |
| SLC25A22 | 454.56 | 2.36E-11 | 11 | 790475-798316 | 9.21E-06 | | |
| SNCA | 26.87 | 9.93E-13 | 4 | 90645250-90759466 | 3.88E-07 | | T |
| TNNT3 | 26.57 | 6.56E-12 | 11 | 1940792-1959936 | 2.56E-06 | | |
| WNT3 | 347.42 | 4.12E-11 | 17 | 44839872-44910520 | 1.61E-05 | | T |
| WNT9B | 23.30 | 4.25E-11 | 17 | 44910567-44964096 | 1.66E-05 | | T |

#### S Table 6: Differential gene expression analysis of candidate genes using PulmonDB data

| Gene | Study | Mean | T statistic | df | P value | Adjusted P value (Bonferroni) | UTMOST effect size in lung | UTMOST P value in lung |
|---|---|---|---|---|---|---|---|---|
| CDHR5 | GSE32537 | 0.086 | 20.69 | 215 | 4.67E-53 | 4.90E-51 | -0.47 | 0.051 |
| LMNTD2 | GSE32537 | -0.097 | -18.04 | 215 | 6.31E-45 | 6.62E-43 | | |
| B4GALNT4 | GSE32537 | -0.071 | -17.44 | 215 | 5.09E-43 | 5.35E-41 | 66.09 | 0.015 |
| SLC25A22 | GSE48149 | -1.023 | -22.63 | 51 | 2.92E-28 | 3.07E-26 | 27.24 | 3.94E-26 |
| HRAS | GSE48149 | -0.325 | -19.20 | 51 | 5.40E-25 | 5.67E-23 | 0.30 | 0.002 |
| B4GALNT4 | GSE38958 | -0.161 | -13.22 | 113 | 1.11E-24 | 1.16E-22 | 66.09 | 0.015 |
| CDHR5 | GSE38958 | -0.113 | -12.42 | 113 | 7.34E-23 | 7.71E-21 | -0.47 | 0.051 |
| B4GALNT4 | GSE48149 | 0.345 | 13.87 | 51 | 6.12E-19 | 6.42E-17 | 66.09 | 0.015 |
| SCT | GSE38958 | -0.146 | -10.26 | 113 | 7.55E-18 | 7.93E-16 | 0.01 | 0.004 |
| SLC25A22 | GSE53845 | 0.171 | 9.34 | 93 | 5.06E-15 | 5.31E-13 | 27.24 | 3.94E-26 |
| B4GALNT4 | GSE53845 | -0.157 | -8.67 | 93 | 1.34E-13 | 1.40E-11 | 66.09 | 0.015 |
| LMNTD2 | GSE38958 | -0.096 | -7.97 | 113 | 1.37E-12 | 1.44E-10 | | |
| LMNTD2 | GSE53845 | -0.084 | -7.86 | 93 | 6.74E-12 | 7.08E-10 | | |
| SLC25A22 | GSE45686 | 0.949 | 9.32 | 37 | 3.05E-11 | 3.20E-09 | 27.24 | 3.94E-26 |
| HRAS | GSE53845 | -0.057 | -7.39 | 93 | 6.29E-11 | 6.60E-09 | 0.30 | 0.002 |
| SLC25A22 | GSE38958 | -0.076 | -7.05 | 113 | 1.53E-10 | 1.60E-08 | 27.24 | 3.94E-26 |
| SCT | GSE32537 | -0.037 | -6.48 | 215 | 6.04E-10 | 6.34E-08 | 0.01 | 0.004 |
| SCT | GSE53845 | -0.079 | -6.45 | 93 | 4.89E-09 | 5.14E-07 | 0.01 | 0.004 |
| SLC25A22 | GSE32537 | -0.044 | -5.90 | 215 | 1.42E-08 | 1.50E-06 | 27.24 | 3.94E-26 |
| B4GALNT4 | GSE21369 | 0.083 | 5.63 | 27 | 5.57E-06 | 0.00058456 | 66.09 | 0.015 |
| POT1 | GSE32537 | 0.038 | 4.43 | 215 | 1.50E-05 | 0.00157869 | -1.48 | 0.231 |
| SLC25A22 | GSE21369 | -0.291 | -5.18 | 27 | 1.88E-05 | 0.00197608 | 27.24 | 3.94E-26 |
| POT1 | GSE48149 | 0.030 | 4.58 | 51 | 2.99E-05 | 0.00314272 | -1.48 | 0.231 |
| HRAS | GSE52463 | -0.444 | -5.61 | 13 | 8.42E-05 | 0.00884564 | 0.30 | 0.002 |
| SCT | GSE21369 | 0.096 | 4.47 | 27 | 0.00012689 | 0.01332393 | 0.01 | 0.004 |
| SLC25A22 | GSE52463 | -0.455 | -4.77 | 13 | 0.00036662 | 0.03849502 | 27.24 | 3.94E-26 |

**S Table 7A: Partitioned heritability results of traits used in MTAG. P values are adjusted using Bonferroni correction**

| IMPACT index | Tissue | Cell Deriv | Trait | Enrichment | Adjusted P value |
|---|---|---|---|---|---|
| 432 | Blood | B cell | AFP | 1.414594 | 3.22E-01 |
| | | | BFP | 1.381341 | 3.59E-01 |
| | | | HC | 1.433832 | 2.08E-01 |
| | | | WBF | 1.387299 | 1.67E-01 |
| | | | IPF | 1.9543399 | 0.215503 |
| 640 | Uterus | Cervix | AFP | 4.683332 | 3.98E-01 |
| | | | BFP | 4.454029 | 3.81E-01 |
| | | | HC | 4.940997 | 2.10E-01 |
| | | | WBF | 4.305172 | 2.16E-01 |
| | | | IPF | 4.7139771 | 0.5111049 |
| 641 | Stemcell | Stem cell | AFP | 2.584577 | 5.74E-02 |
| | | | BFP | 2.44071 | 5.29E-02 |
| | | | HC | 2.621385 | 7.45E-02 |
| | | | WBF | 2.441791 | 2.41E-01 |
| | | | IPF | 0.7777278 | 0.8823101 |
| 646 | Liver | Liver | AFP | 1.4834 | 3.21E-01 |
| | | | BFP | 1.463281 | 1.47E-01 |
| | | | HC | 1.524422 | 1.88E-01 |
| | | | WBF | 1.432233 | 6.59E-02 |
| | | | IPF | 1.2784806 | 0.7912263 |
| 650 | Blood | Myeloid | AFP | 1.646654 | 1.40E-01 |
| | | | BFP | 1.587917 | 1.92E-01 |
| | | | HC | 1.730841 | 6.71E-02 |
| | | | WBF | 1.591209 | 3.90E-01 |
| | | | IPF | 1.9676699 | 0.5111049 |
| 669 | Fibroblast | Lung | AFP | 1.687388 | 1.57E-01 |
| | | | BFP | 1.67792 | 3.02E-01 |
| | | | HC | 1.741407 | 2.37E-01 |
| | | | WBF | 1.608516 | 8.88E-02 |
| | | | IPF | 1.2534855 | 0.8494849 |

**S Table 7B: *MAFK* Partitioned heritability results of IPF after MTAG. P values are adjusted using Bonferroni correction**

| IMPACT index | Tissue | Cell Deriv | Trait for MTAG | Enrichment | Adjusted P value |
|---|---|---|---|---|---|
| 432 | Blood | B cell | AFP | 1.470080161 | 0.002878912 |
| | | | BFP | 1.407362643 | 0.013601266 |
| | | | HC | 1.458294923 | 0.016867131 |
| | | | WBF | 1.426105821 | 0.022188703 |
| 640 | Uterus | Cervix | AFP | 4.638252288 | 0.007389846 |
| | | | BFP | 4.171382438 | 0.028723909 |
| | | | HC | 4.514322749 | 0.014378828 |
| | | | WBF | 3.992695315 | 0.096248279 |
| 641 | Stemcell | Stem cell | AFP | 2.582370731 | 0.00790598 |
| | | | BFP | 2.367606463 | 0.025863991 |
| | | | HC | 2.505947582 | 0.026093808 |
| | | | WBF | 2.340081324 | 0.074596358 |
| 646 | Liver | Liver | AFP | 1.505094586 | 0.001713691 |
| | | | BFP | 1.434992768 | 0.01242485 |
| | | | HC | 1.516976752 | 0.001613822 |
| | | | WBF | 1.419118026 | 0.051209956 |
| 650 | Blood | Myeloid | AFP | 1.679793221 | 0.095859178 |
| | | | BFP | 1.579624259 | 0.52228124 |
| | | | HC | 1.744878055 | 0.094988905 |
| | | | WBF | 1.590247895 | 0.702155452 |
| 669 | Fibroblast | Lung | AFP | 1.737942699 | 4.74E-05 |
| | | | BFP | 1.672887944 | 0.000122439 |
| | | | HC | 1.738980327 | 0.000533098 |
| | | | WBF | 1.617758927 | 0.007663334 |

**S Table 8A:** ***MAFK*** **targeted gene set from ChIP-Atlas**

| Index | Cell type Class | Cell type | Tissue | Tissue diagnosis/lineage |
|---|---|---|---|---|
| SRX150731 | Blood | GM12878 | blood | mesoderm |
| SRX186621 | Blood | GM12878 | blood | mesoderm |
| SRX150370 | Uterus | HeLa | Cervix | Adenocarcinoma |
| SRX150385 | Liver | Hep G2 | Liver | Carcinoma Hepatocellular |
| SRX150689 | Liver | Hep G2 | Liver | Carcinoma Hepatocellular |
| SRX150372 | Pluripotent stem cell | hESC H1 | | |
| SRX150483 | Lung | IMR-90 | Lung | Normal |
| SRX150391 | Blood | K-562 | Blood | Leukemia Chronic Myelogenous |
| SRX150493 | Blood | K-562 | Blood | Leukemia Chronic Myelogenous |
| ERX159255 | Digestive tract | LoVo | Colon | Adenocarcinoma |
| ERX159256 | Digestive tract | LoVo | Colon | Adenocarcinoma |
| SRX360026 | Digestive tract | LoVo | Colon | Adenocarcinoma |
| SRX298273 | Blood | OCI-LY-7 | Blood | Lymphoma B-cell |

**S Table 8B:** ***SMAD2*** **targeted gene set from ChIP-Atlas**

| Index | Cell type Class | Cell type | Tissue | Tissue diagnosis/lineage |
|---|---|---|---|---|
| SRX3847778 | Cardiovascular | Aortic_smooth_muscle_cells | | |
| SRX3847781 | Cardiovascular | Aortic_smooth_muscle_cells | | |
| SRX6476497 | Cardiovascular | Aortic_smooth_muscle_cells | | |
| SRX6476500 | Cardiovascular | Aortic_smooth_muscle_cells | | |
| SRX702089 | Pluripotent stem cell | hESC_derived_ectodermal_cells | | |
| SRX064489 | Pluripotent stem cell | hESC_derived_mesendodermal_cells | | |
| SRX064490 | Pluripotent stem cell | hESC_derived_mesendodermal_cells | | |
| SRX084502 | Pluripotent stem cell | hESC_derived_mesendodermal_cells | | |
| SRX084503 | Pluripotent stem cell | hESC_derived_mesendodermal_cells | | |
| SRX702090 | Pluripotent stem cell | hESC_derived_mesendodermal_cells | | |
| SRX702091 | Pluripotent stem cell | hESC_derived_mesendodermal_cells | | |
| SRX2844311 | Pluripotent stem cell | hESC_H1 | | |
| SRX2844312 | Pluripotent stem cell | hESC_H1 | | |
| SRX064480 | Pluripotent stem cell | hESC_H9 | | |
| SRX064481 | Pluripotent stem cell | hESC_H9 | | |
| SRX084500 | Pluripotent stem cell | hESC_H9 | | |
| SRX084501 | Pluripotent stem cell | hESC_H9 | | |
| SRX702088 | Pluripotent stem cell | hESC_HUES64 | | |
| SRX3592677 | Pluripotent stem cell | hESC_HUES8 | | |
| SRX3592681 | Pluripotent stem cell | hESC_HUES8 | | |
| SRX3592685 | Pluripotent stem cell | hESC_HUES8 | | |
| SRX378126 | Pluripotent stem cell | hESC_WA09 | | |
| SRX378127 | Pluripotent stem cell | hESC_WA09 | | |
| SRX6957406 | Kidney | HGrC1 | | |
| SRX6957412 | Kidney | HGrC1 | | |
| SRX6957413 | Kidney | HGrC1 | | |
| SRX6957414 | Kidney | HGrC1 | | |
| SRX6957415 | Kidney | HGrC1 | | |
| SRX6957436 | Kidney | HGrC1 | | |
| SRX6957437 | Kidney | HGrC1 | | |
| SRX6957486 | Kidney | HGrC1 | | |
| SRX6957491 | Kidney | HGrC1 | | |
| SRX6476489 | Cardiovascular | HUVEC | Umbilical Cord | Normal |
| SRX6476492 | Cardiovascular | HUVEC | Umbilical Cord | Normal |
| SRX6957479 | Gonad | KGN | Ovary | Tumor Granulosa Cell |
| SRX6957483 | Gonad | KGN | Ovary | Tumor Granulosa Cell |
| SRX2245587 | Pancreas | PANC-1 | Pancreas/Duct | Epithelioid Carcinoma |
| SRX2245588 | Pancreas | PANC-1 | Pancreas/Duct | Epithelioid Carcinoma |

**S Table 9A: Enrichment of *MAFK* targets in cell-type-specific differentially expressed genes in IPF single cell data. P values are adjusted using Bonferroni correction**

| Annotation | # genes | Celltype | P value | Adjusted P value (Bonferroni) |
|---|---|---|---|---|
| SRX150483.IMR.90 | 64 | Myofibroblast | 1.89E-50 | 8.45E-48 |
| SRX150385.Hep_G2 | 55 | Macrophage | 3.53E-49 | 1.58E-46 |
| SRX150483.IMR.90 | 54 | Macrophage | 2.95E-48 | 1.32E-45 |
| SRX150385.Hep_G2 | 53 | Macrophage_Alveolar | 2.64E-45 | 1.18E-42 |
| SRX150483.IMR.90 | 53 | Macrophage_Alveolar | 2.64E-45 | 1.18E-42 |
| SRX150385.Hep_G2 | 66 | Ciliated | 2.15E-41 | 9.63E-39 |
| SRX150385.Hep_G2 | 44 | VE_Capillary_B | 2.56E-40 | 1.15E-37 |
| SRX150385.Hep_G2 | 51 | Myofibroblast | 5.61E-40 | 2.51E-37 |
| SRX150483.IMR.90 | 41 | VE_Capillary_B | 1.60E-37 | 7.16E-35 |
| SRX150689.Hep_G2 | 41 | Macrophage | 2.03E-36 | 9.07E-34 |
| SRX150391.K.562 | 41 | Macrophage | 2.03E-36 | 9.07E-34 |
| SRX150483.IMR.90 | 57 | Ciliated | 1.15E-35 | 5.13E-33 |
| SRX150370.HeLa | 40 | Macrophage | 1.61E-35 | 7.20E-33 |
| SRX150483.IMR.90 | 38 | Fibroblast | 4.51E-35 | 2.02E-32 |
| SRX150385.Hep_G2 | 46 | ATII | 5.75E-35 | 2.58E-32 |
| SRX150483.IMR.90 | 44 | ATII | 1.97E-33 | 8.81E-31 |
| SRX150689.Hep_G2 | 38 | Macrophage_Alveolar | 2.60E-32 | 1.17E-29 |
| SRX150391.K.562 | 38 | Macrophage_Alveolar | 2.60E-32 | 1.17E-29 |
| SRX150385.Hep_G2 | 35 | Fibroblast | 2.81E-32 | 1.26E-29 |
| SRX150689.Hep_G2 | 41 | Myofibroblast | 4.67E-32 | 2.09E-29 |
| SRX150370.HeLa | 50 | Ciliated | 2.96E-31 | 1.33E-28 |
| SRX150370.HeLa | 36 | Macrophage_Alveolar | 1.33E-30 | 5.97E-28 |
| SRX150689.Hep_G2 | 32 | VE_Capillary_B | 3.18E-29 | 1.43E-26 |
| SRX150385.Hep_G2 | 31 | ATI | 4.01E-29 | 1.80E-26 |
| SRX150483.IMR.90 | 31 | ATI | 4.01E-29 | 1.80E-26 |
| SRX150689.Hep_G2 | 44 | Ciliated | 1.68E-27 | 7.54E-25 |
| SRX150483.IMR.90 | 19 | ncMonocyte | 1.14E-26 | 5.10E-24 |
| SRX150385.Hep_G2 | 24 | Club | 1.38E-26 | 6.19E-24 |
| SRX150385.Hep_G2 | 20 | cMonocyte | 2.97E-26 | 1.33E-23 |
| SRX150370.HeLa | 28 | VE_Capillary_B | 1.41E-25 | 6.32E-23 |
| SRX150483.IMR.90 | 19 | cMonocyte | 6.13E-25 | 2.75E-22 |
| SRX150391.K.562 | 27 | VE_Capillary_B | 1.14E-24 | 5.11E-22 |
| SRX150689.Hep_G2 | 31 | ATII | 1.44E-23 | 6.47E-21 |
| SRX150370.HeLa | 30 | Myofibroblast | 1.78E-23 | 7.99E-21 |
| SRX150391.K.562 | 30 | Myofibroblast | 1.78E-23 | 7.99E-21 |
| SRX150689.Hep_G2 | 25 | Fibroblast | 4.53E-23 | 2.03E-20 |
| SRX150370.HeLa | 30 | ATII | 8.15E-23 | 3.65E-20 |
| SRX150689.Hep_G2 | 24 | ATI | 1.41E-22 | 6.33E-20 |
| SRX150391.K.562 | 36 | Ciliated | 1.56E-22 | 6.97E-20 |
| SRX150391.K.562 | 29 | ATII | 4.59E-22 | 2.06E-19 |
| SRX150370.HeLa | 23 | Fibroblast | 3.01E-21 | 1.35E-18 |

| Annotation | # genes | Celltype | P value | Adjusted P value (Bonferroni) |
|---|---|---|---|---|
| SRX150385.Hep_G2 | 18 | cDC2 | 3.94E-21 | 1.76E-18 |
| SRX150385.Hep_G2 | 15 | ncMonocyte | 4.64E-21 | 2.08E-18 |
| SRX150483.IMR.90 | 18 | Club | 5.45E-20 | 2.44E-17 |
| SRX150391.K.562 | 21 | ATI | 8.53E-20 | 3.82E-17 |
| SRX150370.HeLa | 15 | cMonocyte | 1.01E-19 | 4.52E-17 |
| SRX150391.K.562 | 15 | cMonocyte | 1.01E-19 | 4.52E-17 |
| SRX150370.HeLa | 14 | ncMonocyte | 1.14E-19 | 5.10E-17 |
| SRX150391.K.562 | 14 | ncMonocyte | 1.14E-19 | 5.10E-17 |
| SRX150385.Hep_G2 | 14 | Lymphatic | 1.82E-19 | 8.14E-17 |
| SRX150483.IMR.90 | 14 | Lymphatic | 1.82E-19 | 8.14E-17 |
| SRX150370.HeLa | 20 | ATI | 7.16E-19 | 3.21E-16 |
| SRX150483.IMR.90 | 16 | cDC2 | 8.07E-19 | 3.62E-16 |
| SRX150391.K.562 | 20 | Fibroblast | 1.59E-18 | 7.11E-16 |
| SRX150385.Hep_G2 | 13 | NK | 1.76E-18 | 7.91E-16 |
| SRX150689.Hep_G2 | 14 | cMonocyte | 1.98E-18 | 8.88E-16 |
| SRX150385.Hep_G2 | 11 | VE_Venous | 4.56E-18 | 2.04E-15 |
| SRX298273.OCI.LY.7 | 21 | Macrophage_Alveolar | 5.92E-18 | 2.65E-15 |
| SRX150372.hESC_H1 | 19 | ATI | 5.98E-18 | 2.68E-15 |
| SRX150372.hESC_H1 | 20 | Macrophage | 7.66E-18 | 3.43E-15 |
| SRX150385.Hep_G2 | 12 | T_Cytotoxic | 1.59E-17 | 7.12E-15 |
| SRX150372.hESC_H1 | 22 | Myofibroblast | 2.57E-17 | 1.15E-14 |
| SRX150689.Hep_G2 | 12 | Lymphatic | 9.82E-17 | 4.40E-14 |
| SRX150370.HeLa | 14 | cDC2 | 1.61E-16 | 7.22E-14 |
| SRX150483.IMR.90 | 10 | VE_Venous | 1.89E-16 | 8.47E-14 |
| SRX150385.Hep_G2 | 11 | T | 4.27E-16 | 1.91E-13 |
| SRX150689.Hep_G2 | 11 | NK | 1.08E-15 | 4.82E-13 |
| SRX150689.Hep_G2 | 14 | Club | 1.21E-15 | 5.43E-13 |
| SRX150689.Hep_G2 | 11 | ncMonocyte | 1.56E-15 | 7.01E-13 |
| SRX150372.hESC_H1 | 18 | Macrophage_Alveolar | 1.85E-15 | 8.28E-13 |
| SRX150689.Hep_G2 | 13 | cDC2 | 2.26E-15 | 1.01E-12 |
| SRX150391.K.562 | 13 | cDC2 | 2.26E-15 | 1.01E-12 |
| SRX150372.hESC_H1 | 20 | ATII | 2.36E-15 | 1.06E-12 |
| SRX298273.OCI.LY.7 | 17 | Macrophage | 3.06E-15 | 1.37E-12 |
| SRX150372.hESC_H1 | 24 | Ciliated | 3.61E-15 | 1.62E-12 |
| SRX150483.IMR.90 | 10 | T | 1.13E-14 | 5.08E-12 |
| SRX150391.K.562 | 10 | T | 1.13E-14 | 5.08E-12 |
| SRX150689.Hep_G2 | 10 | T_Cytotoxic | 1.13E-14 | 5.08E-12 |
| SRX150372.hESC_H1 | 15 | VE_Capillary_B | 6.77E-14 | 3.03E-11 |
| SRX150385.Hep_G2 | 7 | VE_Capillary_A | 1.05E-13 | 4.69E-11 |
| SRX150391.K.562 | 12 | Club | 1.75E-13 | 7.83E-11 |
| SRX150689.Hep_G2 | 9 | T | 2.97E-13 | 1.33E-10 |
| SRX150391.K.562 | 9 | T_Cytotoxic | 2.97E-13 | 1.33E-10 |
| SRX150689.Hep_G2 | 8 | VE_Venous | 3.06E-13 | 1.37E-10 |

| Annotation | # genes | Celltype | P value | Adjusted P value (Bonferroni) |
|---|---|---|---|---|
| SRX150483.IMR.90 | 9 | NK | 6.27E-13 | 2.81E-10 |
| SRX150483.IMR.90 | 5 | DC_Mature | 2.98E-12 | 1.34E-09 |
| SRX150385.Hep_G2 | 7 | B | 7.05E-12 | 3.16E-09 |
| SRX150370.HeLa | 8 | T | 7.69E-12 | 3.44E-09 |
| SRX150370.HeLa | 8 | T_Cytotoxic | 7.69E-12 | 3.44E-09 |
| SRX150391.K.562 | 7 | VE_Venous | 1.20E-11 | 5.36E-09 |
| SRX150370.HeLa | 8 | NK | 1.49E-11 | 6.66E-09 |
| SRX150391.K.562 | 8 | NK | 1.49E-11 | 6.66E-09 |
| SRX298273.OCI.LY.7 | 8 | ncMonocyte | 1.94E-11 | 8.70E-09 |
| SRX150370.HeLa | 10 | Club | 2.47E-11 | 1.10E-08 |
| SRX298273.OCI.LY.7 | 12 | VE_Capillary_B | 3.09E-11 | 1.39E-08 |
| SRX298273.OCI.LY.7 | 14 | Myofibroblast | 3.16E-11 | 1.42E-08 |
| SRX186621.GM12878 | 12 | Macrophage | 6.24E-11 | 2.80E-08 |
| SRX150483.IMR.90 | 7 | T_Cytotoxic | 1.96E-10 | 8.80E-08 |
| SRX360026.LoVo | 11 | VE_Capillary_B | 2.37E-10 | 1.06E-07 |
| SRX298273.OCI.LY.7 | 16 | Ciliated | 2.59E-10 | 1.16E-07 |
| SRX150372.hESC_H1 | 7 | ncMonocyte | 4.40E-10 | 1.97E-07 |
| SRX150370.HeLa | 6 | VE_Venous | 4.59E-10 | 2.06E-07 |
| SRX150689.Hep_G2 | 5 | VE_Capillary_A | 6.40E-10 | 2.87E-07 |
| SRX150483.IMR.90 | 5 | VE_Capillary_A | 6.40E-10 | 2.87E-07 |
| SRX150385.Hep_G2 | 5 | VE_Arterial | 1.22E-09 | 5.48E-07 |
| SRX150372.hESC_H1 | 10 | Fibroblast | 1.49E-09 | 6.69E-07 |
| SRX150372.hESC_H1 | 7 | cMonocyte | 1.74E-09 | 7.81E-07 |
| SRX298273.OCI.LY.7 | 12 | ATII | 1.86E-09 | 8.33E-07 |
| SRX150483.IMR.90 | 4 | VE_Peribronchial | 2.05E-09 | 9.20E-07 |
| SRX360026.LoVo | 10 | Macrophage | 3.23E-09 | 1.45E-06 |
| SRX150372.hESC_H1 | 8 | Club | 3.40E-09 | 1.52E-06 |
| SRX186621.GM12878 | 11 | Myofibroblast | 5.84E-09 | 2.62E-06 |
| SRX186621.GM12878 | 10 | Macrophage_Alveolar | 7.30E-09 | 3.27E-06 |
| SRX298273.OCI.LY.7 | 9 | ATI | 8.15E-09 | 3.65E-06 |
| SRX150370.HeLa | 5 | B | 1.19E-08 | 5.34E-06 |
| SRX150689.Hep_G2 | 5 | B | 1.19E-08 | 5.34E-06 |
| SRX150372.hESC_H1 | 5 | B | 1.19E-08 | 5.34E-06 |
| SRX150483.IMR.90 | 5 | B | 1.19E-08 | 5.34E-06 |
| SRX150391.K.562 | 5 | B | 1.19E-08 | 5.34E-06 |
| SRX150391.K.562 | 6 | Lymphatic | 1.19E-08 | 5.35E-06 |
| SRX186621.GM12878 | 9 | VE_Capillary_B | 1.37E-08 | 6.14E-06 |
| SRX150385.Hep_G2 | 4 | B_Plasma | 1.60E-08 | 7.16E-06 |
| SRX150689.Hep_G2 | 4 | B_Plasma | 1.60E-08 | 7.16E-06 |
| SRX298273.OCI.LY.7 | 4 | B_Plasma | 1.60E-08 | 7.16E-06 |
| SRX298273.OCI.LY.7 | 6 | cMonocyte | 3.19E-08 | 1.43E-05 |
| SRX150391.K.562 | 4 | VE_Capillary_A | 4.73E-08 | 2.12E-05 |
| SRX360026.LoVo | 9 | Macrophage_Alveolar | 4.82E-08 | 2.16E-05 |

| Annotation | # genes | Celltype | P value | Adjusted P value (Bonferroni) |
|---|---|---|---|---|
| SRX360026.LoVo | 9 | Myofibroblast | 1.87E-07 | 8.39E-05 |
| SRX150372.hESC_H1 | 6 | cDC2 | 1.98E-07 | 8.87E-05 |
| SRX298273.OCI.LY.7 | 6 | cDC2 | 1.98E-07 | 8.87E-05 |
| SRX150493.K.562 | 11 | Ciliated | 2.67E-07 | 0.00011983 |
| SRX186621.GM12878 | 9 | ATII | 2.92E-07 | 0.00013062 |
| SRX150385.Hep_G2 | 3 | VE_Peribronchial | 3.42E-07 | 0.00015311 |
| SRX186621.GM12878 | 7 | ATI | 5.24E-07 | 0.00023463 |
| ERX159256.LoVo | 7 | VE_Capillary_B | 7.84E-07 | 0.00035115 |
| SRX150385.Hep_G2 | 3 | Basal | 9.99E-07 | 0.00044755 |
| SRX150689.Hep_G2 | 3 | Basal | 9.99E-07 | 0.00044755 |
| SRX150483.IMR.90 | 3 | Basal | 9.99E-07 | 0.00044755 |
| SRX150391.K.562 | 3 | Basal | 9.99E-07 | 0.00044755 |
| SRX186621.GM12878 | 10 | Ciliated | 1.07E-06 | 0.00047787 |
| SRX186621.GM12878 | 3 | B_Plasma | 1.52E-06 | 0.00068109 |
| SRX150370.HeLa | 3 | B_Plasma | 1.52E-06 | 0.00068109 |
| SRX150372.hESC_H1 | 3 | B_Plasma | 1.52E-06 | 0.00068109 |
| SRX150483.IMR.90 | 3 | B_Plasma | 1.52E-06 | 0.00068109 |
| SRX150372.hESC_H1 | 4 | T | 3.05E-06 | 0.00136475 |
| SRX298273.OCI.LY.7 | 4 | T | 3.05E-06 | 0.00136475 |
| SRX360026.LoVo | 6 | ATI | 4.18E-06 | 0.0018714 |
| SRX150731.GM12878 | 9 | Ciliated | 4.25E-06 | 0.00190286 |
| SRX186621.GM12878 | 4 | ncMonocyte | 4.79E-06 | 0.00214577 |
| SRX360026.LoVo | 4 | ncMonocyte | 4.79E-06 | 0.00214577 |
| SRX150689.Hep_G2 | 3 | VE_Arterial | 4.92E-06 | 0.00220242 |
| SRX150483.IMR.90 | 3 | VE_Arterial | 4.92E-06 | 0.00220242 |
| SRX150391.K.562 | 3 | VE_Arterial | 4.92E-06 | 0.00220242 |
| SRX298273.OCI.LY.7 | 6 | Fibroblast | 5.27E-06 | 0.00236041 |
| SRX150370.HeLa | 4 | Lymphatic | 5.43E-06 | 0.00243475 |
| SRX298273.OCI.LY.7 | 4 | Lymphatic | 5.43E-06 | 0.00243475 |
| SRX150493.K.562 | 6 | Macrophage | 8.35E-06 | 0.00373943 |
| ERX159256.LoVo | 6 | Macrophage | 8.35E-06 | 0.00373943 |
| SRX298273.OCI.LY.7 | 3 | B | 1.86E-05 | 0.00834666 |
| SRX150372.hESC_H1 | 3 | VE_Venous | 2.32E-05 | 0.01038319 |
| SRX150385.Hep_G2 | 2 | DC_Mature | 3.30E-05 | 0.0147914 |
| SRX150391.K.562 | 2 | DC_Mature | 3.30E-05 | 0.0147914 |
| SRX298273.OCI.LY.7 | 2 | DC_Mature | 3.30E-05 | 0.0147914 |
| ERX159256.LoVo | 5 | ATI | 3.32E-05 | 0.01487428 |
| SRX186621.GM12878 | 5 | Fibroblast | 4.03E-05 | 0.01804221 |
| SRX150493.K.562 | 6 | ATII | 4.48E-05 | 0.02008729 |
| SRX360026.LoVo | 6 | ATII | 4.48E-05 | 0.02008729 |
| SRX150370.HeLa | 2 | VE_Peribronchial | 5.25E-05 | 0.02353177 |
| SRX150391.K.562 | 2 | VE_Peribronchial | 5.25E-05 | 0.02353177 |
| SRX298273.OCI.LY.7 | 2 | VE_Peribronchial | 5.25E-05 | 0.02353177 |

| Annotation | # genes | Celltype | P value | Adjusted P value (Bonferroni) |
|---|---|---|---|---|
| ERX159255.LoVo | 5 | Macrophage | 5.91E-05 | 0.02645445 |
| SRX186621.GM12878 | 4 | Club | 6.08E-05 | 0.02725246 |
| SRX360026.LoVo | 4 | Club | 6.08E-05 | 0.02725246 |
| SRX298273.OCI.LY.7 | 4 | Club | 6.08E-05 | 0.02725246 |
| SRX150372.hESC_H1 | 3 | T_Cytotoxic | 7.42E-05 | 0.03323656 |
| SRX360026.LoVo | 3 | T_Cytotoxic | 7.42E-05 | 0.03323656 |
| SRX150493.K.562 | 5 | Macrophage_Alveolar | 8.85E-05 | 0.03963426 |
| ERX159256.LoVo | 5 | Macrophage_Alveolar | 8.85E-05 | 0.03963426 |
| SRX150372.hESC_H1 | 3 | NK | 9.43E-05 | 0.04225825 |
| SRX360026.LoVo | 3 | NK | 9.43E-05 | 0.04225825 |
| ERX159256.LoVo | 3 | ncMonocyte | 0.00010397 | 0.04657728 |
| SRX150370.HeLa | 2 | Basal | 0.00010505 | 0.04706353 |
| SRX150372.hESC_H1 | 2 | Basal | 0.00010505 | 0.04706353 |

**S Table 9B: Enrichment of *SMAD2* targets in cell-type-specific differentially expressed genes in IPF single cell data. P values are adjusted using Bonferroni correction**

| Annotation | # genes | Celltype | P value | Adjusted P value (Bonferroni) |
|---|---|---|---|---|
| SRX6476489.HUVEC | 139 | VE_Capillary_B | 3.14E-138 | 3.92E-135 |
| SRX6476489.HUVEC | 127 | Myofibroblast | 2.22E-104 | 2.77E-101 |
| SRX6476489.HUVEC | 114 | Macrophage_Alveolar | 8.64E-102 | 1.08E-98 |
| SRX6476489.HUVEC | 107 | Macrophage | 1.38E-99 | 1.72E-96 |
| SRX6476489.HUVEC | 122 | ATII | 9.17E-97 | 1.14E-93 |
| SRX6476489.HUVEC | 80 | Fibroblast | 6.86E-76 | 8.56E-73 |
| SRX6476489.HUVEC | 78 | ATI | 2.08E-75 | 2.60E-72 |
| SRX6476489.HUVEC | 104 | Ciliated | 2.88E-66 | 3.60E-63 |
| SRX6476489.HUVEC | 41 | Lymphatic | 1.60E-58 | 1.99E-55 |
| SRX6476489.HUVEC | 42 | cDC2 | 7.53E-50 | 9.40E-47 |
| SRX6476489.HUVEC | 37 | cMonocyte | 2.63E-49 | 3.28E-46 |
| SRX6476489.HUVEC | 33 | T | 5.30E-49 | 6.62E-46 |
| SRX6476489.HUVEC | 34 | ncMonocyte | 1.57E-48 | 1.96E-45 |
| SRX6476489.HUVEC | 38 | Club | 2.40E-42 | 3.00E-39 |
| SRX064481.hESC_H9 | 39 | ATI | 1.04E-36 | 1.30E-33 |
| SRX6476489.HUVEC | 25 | T_Cytotoxic | 1.17E-36 | 1.46E-33 |
| SRX064481.hESC_H9 | 37 | Macrophage | 7.86E-33 | 9.81E-30 |
| SRX6476489.HUVEC | 16 | VE_Capillary_A | 1.02E-31 | 1.27E-28 |
| SRX6476489.HUVEC | 19 | VE_Venous | 2.39E-31 | 2.99E-28 |
| SRX064481.hESC_H9 | 39 | Myofibroblast | 1.74E-30 | 2.17E-27 |
| SRX6476489.HUVEC | 21 | NK | 7.75E-30 | 9.67E-27 |
| SRX3592685.hESC_HUES8 | 33 | Macrophage | 2.89E-29 | 3.60E-26 |
| SRX064481.hESC_H9 | 38 | ATII | 7.40E-29 | 9.24E-26 |
| SRX064481.hESC_H9 | 46 | Ciliated | 9.49E-29 | 1.18E-25 |
| SRX064481.hESC_H9 | 31 | VE_Capillary_B | 2.61E-28 | 3.26E-25 |
| SRX3592685.hESC_HUES8 | 37 | ATII | 4.25E-28 | 5.30E-25 |
| SRX064481.hESC_H9 | 33 | Macrophage_Alveolar | 4.76E-28 | 5.95E-25 |
| SRX3592685.hESC_HUES8 | 33 | Macrophage_Alveolar | 4.76E-28 | 5.95E-25 |
| SRX064481.hESC_H9 | 28 | Fibroblast | 8.14E-26 | 1.02E-22 |
| SRX064481.hESC_H9 | 23 | Club | 1.76E-25 | 2.20E-22 |
| SRX3592685.hESC_HUES8 | 27 | ATI | 2.26E-25 | 2.82E-22 |
| SRX6476492.HUVEC | 29 | Macrophage_Alveolar | 1.16E-24 | 1.44E-21 |
| SRX6476489.HUVEC | 13 | VE_Arterial | 1.17E-24 | 1.46E-21 |
| SRX3592685.hESC_HUES8 | 31 | Myofibroblast | 3.00E-24 | 3.74E-21 |
| SRX6476492.HUVEC | 26 | Macrophage | 4.39E-23 | 5.47E-20 |
| SRX3592685.hESC_HUES8 | 24 | VE_Capillary_B | 5.90E-22 | 7.37E-19 |
| SRX2844312.hESC_H1 | 22 | ATI | 1.01E-20 | 1.26E-17 |
| SRX3592685.hESC_HUES8 | 33 | Ciliated | 1.10E-20 | 1.38E-17 |
| SRX2844312.hESC_H1 | 24 | ATII | 2.51E-18 | 3.13E-15 |
| SRX3592681.hESC_HUES8 | 24 | ATII | 2.51E-18 | 3.13E-15 |
| SRX6957437.HGrC1 | 19 | Fibroblast | 1.27E-17 | 1.59E-14 |
| SRX6476492.HUVEC | 19 | VE_Capillary_B | 1.83E-17 | 2.29E-14 |
| SRX2844312.hESC_H1 | 22 | Myofibroblast | 2.57E-17 | 3.20E-14 |
| SRX6476492.HUVEC | 22 | Myofibroblast | 2.57E-17 | 3.20E-14 |

| Annotation | # genes | Celltype | P value | Adjusted P value (Bonferroni) |
|---|---|---|---|---|
| SRX064481.hESC_H9 | 13 | cMonocyte | 3.86E-17 | 4.82E-14 |
| SRX2844312.hESC_H1 | 18 | VE_Capillary_B | 1.44E-16 | 1.79E-13 |
| SRX6957437.HGrC1 | 21 | Myofibroblast | 1.49E-16 | 1.86E-13 |
| SRX064481.hESC_H9 | 14 | cDC2 | 1.61E-16 | 2.01E-13 |
| SRX6476492.HUVEC | 21 | ATII | 4.27E-16 | 5.33E-13 |
| SRX064481.hESC_H9 | 11 | Lymphatic | 2.24E-15 | 2.80E-12 |
| SRX3592685.hESC_HUES8 | 11 | Lymphatic | 2.24E-15 | 2.80E-12 |
| SRX6476489.HUVEC | 9 | B | 3.84E-15 | 4.79E-12 |
| SRX6957437.HGrC1 | 16 | VE_Capillary_B | 8.73E-15 | 1.09E-11 |
| SRX3592685.hESC_HUES8 | 10 | T | 1.13E-14 | 1.42E-11 |
| SRX3592677.hESC_HUES8 | 19 | ATII | 1.30E-14 | 1.62E-11 |
| SRX6476489.HUVEC | 7 | B_Plasma | 1.38E-14 | 1.72E-11 |
| SRX3592685.hESC_HUES8 | 13 | Club | 1.46E-14 | 1.82E-11 |
| SRX6476492.HUVEC | 23 | Ciliated | 1.47E-14 | 1.83E-11 |
| SRX6957437.HGrC1 | 15 | ATI | 2.81E-14 | 3.51E-11 |
| SRX064481.hESC_H9 | 10 | ncMonocyte | 3.66E-14 | 4.57E-11 |
| SRX3592685.hESC_HUES8 | 15 | Fibroblast | 5.08E-14 | 6.34E-11 |
| SRX6476492.HUVEC | 15 | Fibroblast | 5.08E-14 | 6.34E-11 |
| SRX2844312.hESC_H1 | 15 | Macrophage | 1.64E-13 | 2.04E-10 |
| SRX3592681.hESC_HUES8 | 17 | Myofibroblast | 1.67E-13 | 2.09E-10 |
| SRX3592685.hESC_HUES8 | 10 | cMonocyte | 2.70E-13 | 3.37E-10 |
| SRX6476492.HUVEC | 10 | cMonocyte | 2.70E-13 | 3.37E-10 |
| SRX2844312.hESC_H1 | 9 | T | 2.97E-13 | 3.71E-10 |
| SRX064481.hESC_H9 | 9 | T | 2.97E-13 | 3.71E-10 |
| SRX064481.hESC_H9 | 9 | T_Cytotoxic | 2.97E-13 | 3.71E-10 |
| SRX6957437.HGrC1 | 17 | ATII | 3.90E-13 | 4.86E-10 |
| SRX2844312.hESC_H1 | 14 | Fibroblast | 4.00E-13 | 5.00E-10 |
| SRX3592685.hESC_HUES8 | 9 | ncMonocyte | 8.48E-13 | 1.06E-09 |
| SRX3592681.hESC_HUES8 | 14 | Macrophage | 1.19E-12 | 1.49E-09 |
| SRX3592677.hESC_HUES8 | 13 | ATI | 1.89E-12 | 2.36E-09 |
| SRX3592681.hESC_HUES8 | 13 | Fibroblast | 3.15E-12 | 3.93E-09 |
| SRX064480.hESC_H9 | 13 | VE_Capillary_B | 4.03E-12 | 5.03E-09 |
| SRX3592685.hESC_HUES8 | 10 | cDC2 | 5.94E-12 | 7.42E-09 |
| SRX6476489.HUVEC | 5 | VE_Peribronchial | 1.13E-11 | 1.41E-08 |
| SRX3592681.hESC_HUES8 | 12 | VE_Capillary_B | 3.09E-11 | 3.86E-08 |
| SRX3592677.hESC_HUES8 | 14 | Myofibroblast | 3.16E-11 | 3.94E-08 |
| SRX3592677.hESC_HUES8 | 12 | Macrophage | 6.24E-11 | 7.79E-08 |
| SRX6957437.HGrC1 | 17 | Ciliated | 6.44E-11 | 8.04E-08 |
| SRX064480.hESC_H9 | 11 | ATI | 1.25E-10 | 1.56E-07 |
| SRX3592677.hESC_HUES8 | 11 | Fibroblast | 1.92E-10 | 2.40E-07 |
| SRX064480.hESC_H9 | 7 | T | 1.96E-10 | 2.45E-07 |
| SRX3592681.hESC_HUES8 | 7 | T | 1.96E-10 | 2.45E-07 |
| SRX3592685.hESC_HUES8 | 7 | T_Cytotoxic | 1.96E-10 | 2.45E-07 |
| SRX2844312.hESC_H1 | 16 | Ciliated | 2.59E-10 | 3.24E-07 |
| SRX064481.hESC_H9 | 7 | NK | 3.49E-10 | 4.35E-07 |
| SRX3592685.hESC_HUES8 | 7 | NK | 3.49E-10 | 4.35E-07 |
| SRX3592681.hESC_HUES8 | 7 | ncMonocyte | 4.40E-10 | 5.49E-07 |

| Annotation | # genes | Celltype | P value | Adjusted P value (Bonferroni) |
|---|---|---|---|---|
| SRX6476492.HUVEC | 7 | ncMonocyte | 4.40E-10 | 5.49E-07 |
| SRX064480.hESC_H9 | 11 | Macrophage | 4.50E-10 | 5.61E-07 |
| SRX6957437.HGrC1 | 6 | VE_Venous | 4.59E-10 | 5.73E-07 |
| SRX2844312.hESC_H1 | 7 | Lymphatic | 5.50E-10 | 6.87E-07 |
| SRX6476489.HUVEC | 4 | DC_Mature | 7.45E-10 | 9.29E-07 |
| SRX3592681.hESC_HUES8 | 10 | ATI | 1.01E-09 | 1.26E-06 |
| SRX6476492.HUVEC | 10 | ATI | 1.01E-09 | 1.26E-06 |
| SRX064480.hESC_H9 | 12 | Myofibroblast | 1.03E-09 | 1.28E-06 |
| SRX3592681.hESC_HUES8 | 15 | Ciliated | 1.04E-09 | 1.30E-06 |
| SRX2844312.hESC_H1 | 11 | Macrophage_Alveolar | 1.10E-09 | 1.38E-06 |
| SRX6957437.HGrC1 | 5 | VE_Arterial | 1.22E-09 | 1.53E-06 |
| SRX3592677.hESC_HUES8 | 10 | VE_Capillary_B | 1.80E-09 | 2.25E-06 |
| SRX6957437.HGrC1 | 10 | Macrophage | 3.23E-09 | 4.03E-06 |
| SRX2844312.hESC_H1 | 8 | Club | 3.40E-09 | 4.25E-06 |
| SRX3592677.hESC_HUES8 | 6 | T | 4.96E-09 | 6.19E-06 |
| SRX6957437.HGrC1 | 6 | T | 4.96E-09 | 6.19E-06 |
| SRX6476492.HUVEC | 6 | T | 4.96E-09 | 6.19E-06 |
| SRX2844312.hESC_H1 | 6 | T_Cytotoxic | 4.96E-09 | 6.19E-06 |
| SRX3592681.hESC_HUES8 | 6 | T_Cytotoxic | 4.96E-09 | 6.19E-06 |
| SRX6476492.HUVEC | 6 | T_Cytotoxic | 4.96E-09 | 6.19E-06 |
| SRX6957437.HGrC1 | 10 | Macrophage_Alveolar | 7.30E-09 | 9.12E-06 |
| SRX3592681.hESC_HUES8 | 6 | NK | 8.08E-09 | 1.01E-05 |
| SRX6476492.HUVEC | 6 | NK | 8.08E-09 | 1.01E-05 |
| SRX3592681.hESC_HUES8 | 6 | Lymphatic | 1.19E-08 | 1.49E-05 |
| SRX064480.hESC_H9 | 13 | Ciliated | 1.67E-08 | 2.09E-05 |
| SRX2844312.hESC_H1 | 5 | VE_Venous | 1.73E-08 | 2.16E-05 |
| SRX064481.hESC_H9 | 5 | VE_Venous | 1.73E-08 | 2.16E-05 |
| SRX3592677.hESC_HUES8 | 6 | cMonocyte | 3.19E-08 | 3.98E-05 |
| SRX3592681.hESC_HUES8 | 6 | cMonocyte | 3.19E-08 | 3.98E-05 |
| SRX6476492.HUVEC | 7 | Club | 3.97E-08 | 4.95E-05 |
| SRX064481.hESC_H9 | 4 | VE_Capillary_A | 4.73E-08 | 5.91E-05 |
| SRX6476489.HUVEC | 3 | Goblet | 6.31E-08 | 7.87E-05 |
| SRX3592677.hESC_HUES8 | 12 | Ciliated | 6.70E-08 | 8.36E-05 |
| SRX064480.hESC_H9 | 8 | Fibroblast | 8.93E-08 | 0.00011142 |
| SRX6957437.HGrC1 | 5 | T_Cytotoxic | 1.24E-07 | 0.00015428 |
| SRX6476489.HUVEC | 3 | cDC1 | 1.24E-07 | 0.00015467 |
| SRX2844312.hESC_H1 | 5 | NK | 1.85E-07 | 0.0002313 |
| SRX3592677.hESC_HUES8 | 5 | NK | 1.85E-07 | 0.0002313 |
| SRX3592681.hESC_HUES8 | 6 | cDC2 | 1.98E-07 | 0.00024703 |
| SRX6476492.HUVEC | 6 | cDC2 | 1.98E-07 | 0.00024703 |
| SRX2844312.hESC_H1 | 5 | ncMonocyte | 2.18E-07 | 0.00027252 |
| SRX064480.hESC_H9 | 5 | ncMonocyte | 2.18E-07 | 0.00027252 |
| SRX6476492.HUVEC | 5 | Lymphatic | 2.56E-07 | 0.00031942 |
| SRX064480.hESC_H9 | 9 | ATII | 2.92E-07 | 0.00036388 |
| SRX064480.hESC_H9 | 8 | Macrophage_Alveolar | 3.17E-07 | 0.00039512 |
| SRX3592681.hESC_HUES8 | 8 | Macrophage_Alveolar | 3.17E-07 | 0.00039512 |

| Annotation | # genes | Celltype | P value | Adjusted P value (Bonferroni) |
| --- | --- | --- | --- | --- |
| SRX2844312.hESC_H1 | 5 | cMonocyte | 5.78E-07 | 0.00072187 |
| SRX064480.hESC_H9 | 5 | cMonocyte | 5.78E-07 | 0.00072187 |
| SRX3592685.hESC_HUES8 | 4 | VE_Venous | 6.38E-07 | 0.00079662 |
| SRX064490.hESC_derived_mesendodermal_cells | 7 | VE_Capillary_B | 7.84E-07 | 0.0009782 |
| SRX6476489.HUVEC | 3 | T_Regulatory | 1.16E-06 | 0.0014436 |
| SRX2844311.hESC_H1 | 7 | Macrophage | 1.18E-06 | 0.00146799 |
| SRX2844311.hESC_H1 | 7 | Macrophage_Alveolar | 2.08E-06 | 0.00259036 |
| SRX3592677.hESC_HUES8 | 7 | Macrophage_Alveolar | 2.08E-06 | 0.00259036 |
| SRX2844312.hESC_H1 | 5 | cDC2 | 2.63E-06 | 0.0032855 |
| SRX064480.hESC_H9 | 5 | cDC2 | 2.63E-06 | 0.0032855 |
| SRX6957437.HGrC1 | 5 | cDC2 | 2.63E-06 | 0.0032855 |
| SRX064480.hESC_H9 | 4 | T_Cytotoxic | 3.05E-06 | 0.00380179 |
| SRX3592677.hESC_HUES8 | 4 | T_Cytotoxic | 3.05E-06 | 0.00380179 |
| SRX6957437.HGrC1 | 3 | VE_Capillary_A | 3.38E-06 | 0.00421364 |
| SRX2844311.hESC_H1 | 6 | ATI | 4.18E-06 | 0.00521319 |
| SRX6957437.HGrC1 | 4 | NK | 4.20E-06 | 0.00524638 |
| SRX3592677.hESC_HUES8 | 4 | ncMonocyte | 4.79E-06 | 0.00597751 |
| SRX6957491.HGrC1 | 3 | VE_Arterial | 4.92E-06 | 0.00613532 |
| SRX3592677.hESC_HUES8 | 5 | Club | 5.30E-06 | 0.00661806 |
| SRX3592681.hESC_HUES8 | 5 | Club | 5.30E-06 | 0.00661806 |
| SRX6957437.HGrC1 | 5 | Club | 5.30E-06 | 0.00661806 |
| SRX6957437.HGrC1 | 4 | Lymphatic | 5.43E-06 | 0.00678252 |
| SRX2844311.hESC_H1 | 6 | VE_Capillary_B | 5.90E-06 | 0.00736051 |
| SRX2844311.hESC_H1 | 7 | Myofibroblast | 5.95E-06 | 0.00742512 |
| SRX064490.hESC_derived_mesendodermal_cells | 6 | Macrophage | 8.35E-06 | 0.01041697 |
| SRX6957437.HGrC1 | 2 | Goblet | 1.80E-05 | 0.02247524 |
| SRX064481.hESC_H9 | 3 | B | 1.86E-05 | 0.02325141 |
| SRX3592677.hESC_HUES8 | 3 | VE_Venous | 2.32E-05 | 0.02892459 |
| SRX064490.hESC_derived_mesendodermal_cells | 6 | Myofibroblast | 3.34E-05 | 0.04170616 |

**S Table 10: Study information of GWAS summary statistics**

| Trait | Abbrev | Sample size | Reference |
|---|---|---|---|
| Alcoholism | DrnkWk | 537349 | https://www.nature.com/articles/s41588-018-0307-5 |
| Alzheimer's Disease | AD | 63926 | https://www.niagads.org/igap-rv-summary-stats-kunkle-p-value-data |
| Anorexia Nervosa | AN | 72517 | https://www.nature.com/articles/s41588-019-0439-2 |
| Anxiety Disorder | ADs | 31890 | https://jamanetwork.com/journals/jamapsychiatry/fullarticle/2733149 |
| Asthma | asthma | 127669 | https://www.nature.com/articles/s41588-017-0014-7 |
| Bipolar Disorder | BD | 51710 | https://www.nature.com/articles/s41588-019-0397-8 |
| Body Mass Index | BMI | 795640 | https://academic.oup.com/hmg/article/27/20/3641/5067845 |
| Breast Cancer | BC | 228951 | https://www.nature.com/articles/nature24284 |
| Chronic Kidney Disease | CKD | 118147 | https://www.nature.com/articles/ncomms10023 |
| Crohn's Disease | Crohn | 40266 | https://www.nature.com/articles/ng.3760 |
| Epilepsy | epilepsy | 34853 | https://www.sciencedirect.com/science/article/pii/S1474442214701711?via%3Dihub |
| HDL Cholesterol | HDL | 99900 | https://www.nature.com/articles/nature09270 |
| Height | Height | 709706 | https://academic.oup.com/hmg/article/27/20/3641/5067845 |
| Inflammatory Bowel Disease | IBD | 59957 | https://www.nature.com/articles/ng.3760 |
| Insomnia | Insomnia | 113006 | https://www.nature.com/articles/ng.3888 |
| Ischaemic Stroke | IS | 29633 | https://n.neurology.org/content/86/13/1217.long |
| LDL Cholesterol | LDL | 95454 | https://www.nature.com/articles/nature09270 |
| Lung Cancer | LC | 85716 | https://pubmed.ncbi.nlm.nih.gov/28604730/ |
| Major Depressive Disorder | MDD | 500199 | http://dx.doi.org/10.1038/s41593-018-0326-7 |
| Neuroticism | NSM | 170911 | https://www.nature.com/articles/ng.3552 |
| Parkinson's Disease | PD | 482730 | https://www.biorxiv.org/content/10.1101/388165v3 |
| Primary Biliary Cirrhosis | PBC | 13239 | https://www.nature.com/articles/ncomms9019 |
| Resting Heart Rate | RHR | 134251 | http://dx.doi.org/10.1038/ng.3708 |
| Rheumatoid Arthritis | RA | 58284 | https://www.nature.com/articles/nature12873 |
| Schizophrenia | SCZ | 105318 | https://www.nature.com/articles/s41588-018-0059-2 |
| Sleep Duration | SD | 127573 | https://journals.plos.org/plosgenetics/article?id=10.1371/journal.pgen.1006125 |
| Systemic Lupus Erythematosus | SLE | 14267 | http://dx.doi.org/10.1038/ng.3434 |
| Total Cholesterol | TC | 100184 | https://www.nature.com/articles/nature09270 |
| Triglycerides | TG | 96598 | https://www.nature.com/articles/nature09270 |
| Type-II Diabetes | T2D | 159208 | http://diabetes.diabetesjournals.org/cgi/pmidlookup?view=long&pmid=28566273 |
| Ulcerative Colitis | UC | 45975 | https://www.nature.com/articles/ng.3760 |
| Monocyte count | Monocyte | 131305 | https://www.sciencedirect.com/science/article/pii/S0092867416314635?via%3Dihub |
